# Blinatumomab Trimer Formation: Insights From A Mechanistic PKPD Model Into The Implications For Switching From Infusion To Subcutaneous Dosing Regimen

**DOI:** 10.1101/2024.03.11.24304117

**Authors:** Georgi I. Kapitanov, Sarah A. Head, David Flowers, Joshua F. Apgar, Joshuaine Grant

## Abstract

Blinatumomab is a bispecific T-cell engager (BiTE) that binds to CD3 on T cells and CD19 on B cells. It has been approved for use in B-cell acute lymphoblastic leukemia (B-ALL) with a regimen that requires continuous infusion (cIV) for four weeks per treatment cycle. It is currently in clinical trials for Non-Hodgkin lymphoma (NHL) with cIV administration. Recently, there have been studies investigating dose-response after subcutaneous (SC) dosing in B-ALL and in NHL to determine whether this more convenient method of delivery would have a similar efficacy/safety profile as continuous infusion. We constructed mechanistic PKPD models of blinatumomab activity in B-ALL and NHL patients, investigating the amount of CD3:blinatumomab:CD19 trimers the drug forms at different dosing administrations and regimens.

The modeling and analysis demonstrate that the explored SC doses in B-ALL and NHL achieve similar trimer numbers as the cIV doses in those indications. We further simulated various subcutaneous dosing regimens, and identified conditions where trimer formation dynamics are similar between constant infusion and subcutaneous dosing. Based on the model results, subcutaneous dosing is a viable and convenient strategy for blinatumomab and is projected to result in similar trimer numbers as constant infusion.

## Introduction

Current treatments for cancers with B-cell derived malignancy, such as Non-Hodgkin lymphoma, show improved outcomes for some patients, but there are still many patients experiencing relapse, refractory disease or resistance. Blinatumomab (BLINCYTO™, Amgen,Thousand Oaks, CA) is a bispecific T-cell engager (BiTE) that binds to CD3 on T cells and CD19 on B cells, resulting in T-cell mediated lysis of the B-cells. The lysis is mediated in part by the trimeric complex (trimer) that is formed through the binding of blinatumomab to both CD3 and CD19. The trimer promotes the formation of an immunological synapse between the T- and B-cells, leading to T-cell activation, T-cell proliferation and B-cell lysis. Blinatumomab has been approved for use by the United States Food and Drug Administration (FDA) for adult Philadelphia chromosome negative relapsed or refractory (R/R) B cell progenitor acute lymphoblastic leukemia (B-ALL) in 2014, and for R/R Philadelphia chromosome-positive B-ALL in 2017. Blinatumomab was also approved for the treatment of minimal residual disease (MRD) in B-ALL patients in 2018.

The half-life of blinatumomab is only 2-3 hours, requiring it to be administered through continuous intravenous infusion (cIV) for four weeks per treatment cycle ^1–4^. It is currently in clinical trials for Non-Hodgkin lymphoma (NHL) with cIV administration ^5^. Recently, there have been studies investigating dose-response after subcutaneous (SC) dosing (in B-ALL ^6^ and in NHL ^7^) to evaluate whether this more convenient method of delivery would have a similar efficacy/safety profile as continuous infusion.

While trimer formation is challenging to measure experimentally, there are several examples where mathematical modeling and simulation have assisted in understanding the complexities of TCE molecules and informing quantitative decision making ^8–14^. In this work, literature and available clinical data are used to develop a PKPD model of blinatumomab in B-ALL and NHL. The model was then used to simulate blinatumomab PK and predict trimer formation in the bone marrow in the case of B-ALL or in the tumor environment in NHL patients, both after continuous infusion and subcutaneous administration. The predicted efficacious doses were then compared to those currently being evaluated in the clinic. The model readout used for this comparison is the trimers formed between T cells and cancer cells, since it has been hypothesized that this is the primary driver of pharmacological effect ^8^.

## Methods

### Model Description and Parameterization

We constructed mechanistic PKPD models of B-ALL and NHL that describe blinatumomab PK in circulation, exposure at the site of action (the bone marrow for B-ALL and solid tumor for NHL) and peripheral tissue, engagement with CD3 on T cells and CD19 on normal and malignant B cells, and the amount of trimers the TCE forms at different dosing administrations and regimens. The different components of the model are described below and the Model diagram is presented in Figure 1.

**Figure 1.**
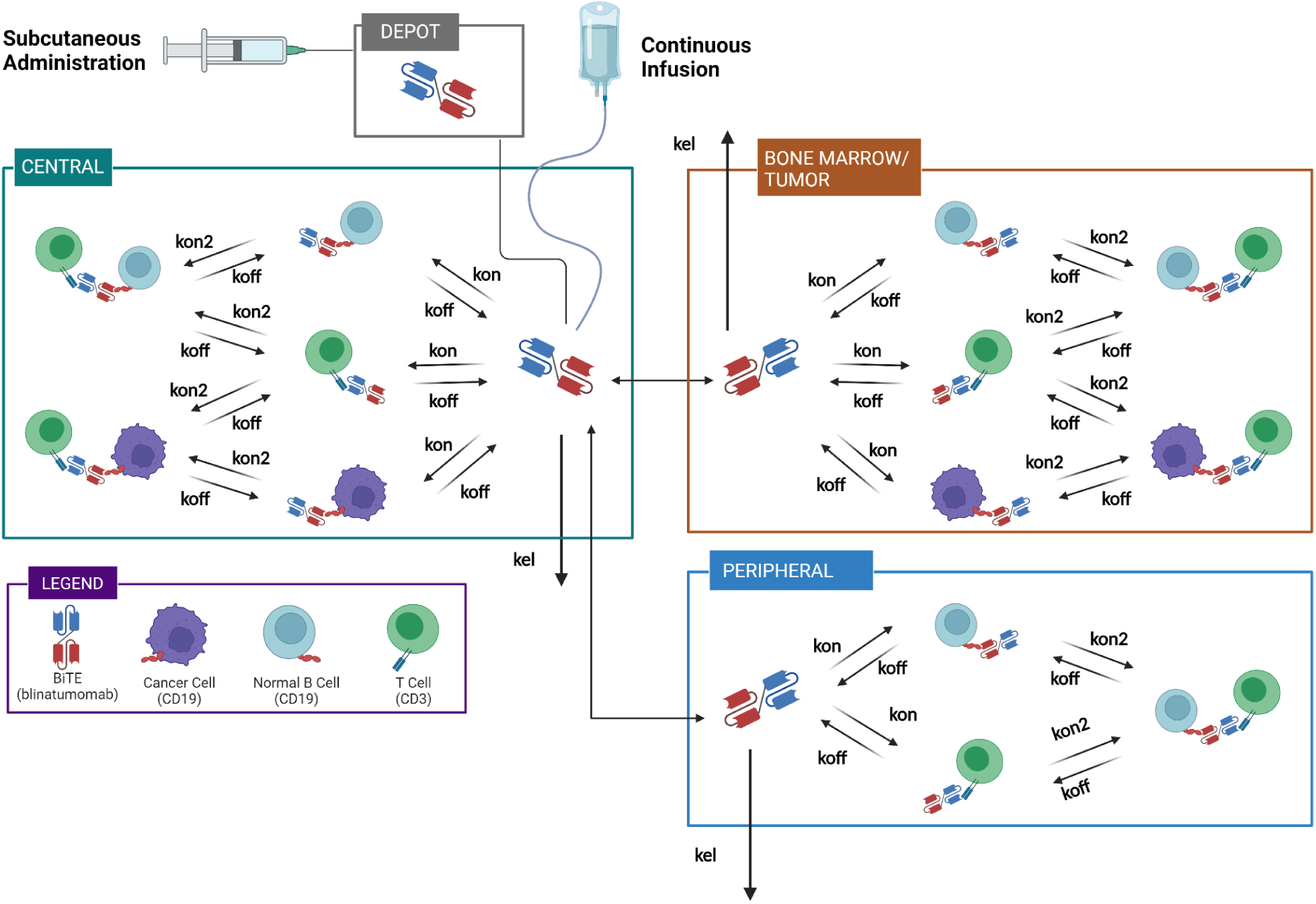
Blinatumomab model diagram. The model includes central, peripheral, and a site of action (bone marrow in B-ALL and tumor in NHL) compartments. CD3 is expressed on T cells, CD19 is expressed on both cancer and normal B cells. Blinatumomab can bind to both receptors on each cell with a kon on-rate and cross-link and form trimers with a kon2 on-rate. The drug can distribute between the central compartment and the other compartments and is eliminated in each compartment. After continuous infusion, blinatumomab enters the central compartment directly. After subcutaneous administration, blinatumomab is absorbed into the central compartment from a depot compartment. Figure created with BioRender.com.

#### Drug administration, distribution, and elimination

Blinatumomab is administered either through continuous intravenous (cIV) infusion or subcutaneously. When cIV administration is applied, blinatumomab enters the central compartment at the clinical reported infusion rate. When administered subcutaneously, blinatumomab is absorbed from a depot compartment, into the central compartment with a rate constant (`kabs`), determined by its half-life (`Thalf_abs_day`), using an applied bioavailability (`fbio`). The drug is assumed to eliminate with the same rate constant (`kel`) in all compartments. Bidirectional distribution from the central to peripheral or central to tumor compartments occurs using blinatumomab distribution parameters `Pdist`, which is the partition coefficient between plasma and the relevant tissue, and `Tdist`, which is the half-life of distribution to that compartment.

The first order drug distribution parameters (k12 and k21, k13 and k31) are calculated from Pdist (partition coefficient between plasma and tissue/tumor) and Tdist (half-life of distribution) through the following algebraic relationship:

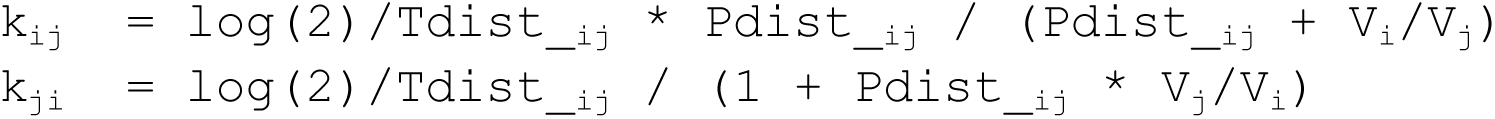

where `i` and `j` are the compartment indexes - 1 for central, 2 for peripheral, 3 for the site of action (tumor or bone marrow). It is assumed that there is no direct distribution between the peripheral compartment and the site of action.

#### CD19 and CD3 protein dynamics

CD19 and CD3 internalization (equivalent to degradation) are included in the model. Synthesis is a zeroth order process, parameterized to ensure constant total (free and bound) protein levels for both targets in each compartment. Internalization is modeled as a single first-order process. The blinatumomab:target complex is assumed to internalize at the rate of the target, while the trimer complex can internalize with either target, releasing the other target in the process to maintain the constant total target level. For example, internalization of the CD3:blinatumomab:CD19 trimer complex through internalization of CD3 results in degradation of CD3 and blinatumomab but returns free CD19. Cellular movement between compartments is not modeled, hence there is no distribution of the targets between compartments.

The initial steady state concentration of CD19 and CD3 was informed from literature estimates of numbers of CD19+ or CD3+ cells, measured receptor expression per cell for both cancer cells and normal B cells, and physiological parameters such as compartment volumes. The cancer cell numbers were determined from clinical data from NHL and B-ALL patients, either by direct cell counts or inferred through the data. These values were selected to represent nominal NHL and B-ALL patients with moderate disease progression. Off-tumor cell counts were obtained from standard values in the literature. For CD19 receptor densities, a representative value was chosen from quantification of receptors per cell that was reported in the literature. The turnover rates of both CD3 and CD19 were obtained from the literature. For all parameter values and references see Tables <u>S1</u> and <u>S2</u>.

#### Binding of blinatumomab to CD3 and CD19, cross-linking

Blinatumomab can bind to either CD3 or CD19 on T cells and B cells, respectively, after which a second binding step, in which blinatumomab binds the target on the other cell type, is modeled. The second binding (cross-linking) creates a CD3:blinatumomab:CD19 trimeric molecule (“trimer”). The one-step binding of blinatumomab to either target is modeled through a traditional concentration-dependent reversible binding step. The second binding step results in trimer formation, is modeled as occurring on the cell surface, and is dependent on the density (molecule per cell surface area) of the blinatumomab:CD3 complex on T cells and the CD19 density on B cells or blinatumomab:CD19 complex on B cells, and the CD3 density on T cells. A detailed description and discussion of the second binding step as density-dependent, and the advantages of this approach in modeling T-cell engagers, can be read in Flowers, D. *et al.*^15^.

All binding reactions are reversible and described with association and dissociation rate constants. The association rate parameter (`kon`) was assumed to be the same for all binding interactions, based on reported protein-protein interaction kinetics ^16^. Blinatumomab’s KDs to CD3 and CD19 are 260 nM and 1.49 nM, respectively, and were taken from the literature ^17^.

The cross-linking association rate, `kon2`, was assumed to be 1000 dm^2^/nmol/s, a typical value based on unpublished internal research experience. The effect of this value on the final dose projections is explored through sensitivity analysis (see Results).

#### B cell depletion

In addition to the basal turnover of CD19 on normal B cells, the models include explicit normal B cell killing. Including B-cell depletion is done to capture potential effects of TMDD. Cancer cells, for the purpose of the model, are not killed. The killing function is trimer-dependent and defines a rate at which B cells are removed from the compartment in which they reside. The same removal rate is applied to all CD19-dependent molecules on normal B cells (free CD19, blinatumomab:CD19 complex, and trimers) and it is assumed that the CD19 associated with that complex is removed from the system, while other species are dissociated from that complex. For example, a blinatumomab:CD19 complex to which the killing rate is applied returns unbound-blinatumomab, while a trimer to which the killing rate is applied will return a blinatumomab:CD3 complex.

The function that defines B-cell depletion, f_kill, has the form:

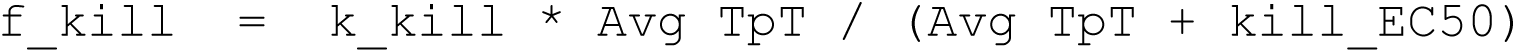

where `k_kill` is the maximum killing velocity, `Avg TpT` is the running average Trimers per T cell (TpT) in the relevant model compartment, and `kill_EC50` is the `Avg TpT` that results in half-max depletion. `kill_EC50` was fixed at 100, meaning that an Avg Tpt of 100 will result in half-max depletion of normal B cells; the effect of this value on the final dose projections is explored through sensitivity analysis (see Results). `k_kill` was determined by a literature search on the effect of blinatumomab on B-cell depletion. Depletion half-life estimates from the literature ranged from less than 1 h to up to 6 h^5,18–20^. Based on these observations, the maximum killing half-life was fixed at 1 h, which resulted in `k_kill` = log(2)/1 h = 2e-4 1/s.

The parameters, their descriptions, along with the assumptions and appropriate references are described in detail in Table S1 for B-ALL and Table S2 for NHL.

#### Analysis Workflow

All blinatumomab parameters besides the absorption half-life after SC dosing (Thalf_abs_day) were fixed to literature values (See <u>Table S1</u> and Table <u>S2</u>). The absorption half-life was estimated from PK data from a blinatumomab NHL study with SC administration ^7^. The data used for model calibration and verification were digitized from published plots<u>(Rossi et al. 2021;</u> <u>Sánchez et al. 2021)</u>. The predicted PK was then compared against reported clinical steady-state concentration levels achieved after continuous infusion of blinatumomab ^5^, as well as clinical PK data from B-ALL patients after SC administration ^6^. The cIV NHL study doses were reported in ug/m^2^/day but in subsequent NHL studies, blinatumomab was dosed in ug. The model input was in ug doses only, which were adjusted accordingly for the cIV study for an assumed typical 70 kg, 175 cm patient.

To compare cIV and SC dosing, a metric equating average trimer per T cell (TpT) for trimers connecting cancer and T cells at the site of action (tumor for NHL and bone marrow for B-ALL) was used. The average TpT was calculated by dividing the AUC of trimers per T cell by the time point of interest. Average trimers per T cell at day 28 at cIV clinical doses of interest (28 ug/day for B-ALL and 60 ug/m^2^/day for NHL) were projected by the model. A scan of doses with SC administration to match the number of trimers achieved at the cIV doses was performed with QD and Q2D regimens, in 5 ug increments.

The models were implemented using the Applied BioMath QSP Notebook v2022.6.4 and the text file of the model reactions for the B-ALL model is shared in Supplemental Materials 3.

## Results

The blinatumomab absorption half-life after SC administration was estimated at 0.527 days. The simulated PK profiles vs data in the NHL clinical trial can be seen in Figure 2 and the goodness-of-fit projected vs observed values can be seen in <u>Figure S1</u>.

**Figure 2.**
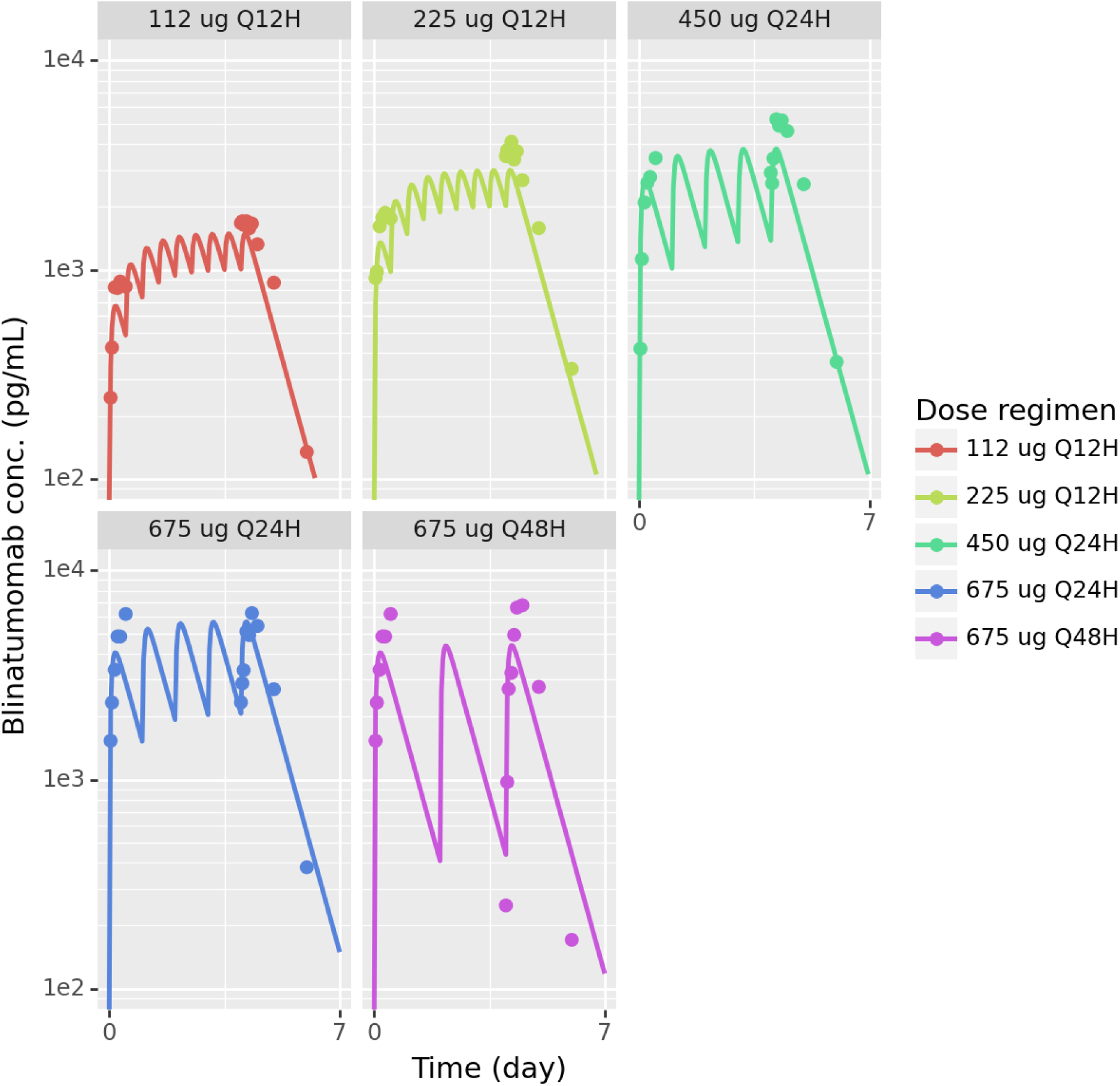
Blinatumomab PK calibration to NHL SC study ^7^. All parameters besides the absorption half-life were taken from the literature.

Next, steady-state concentrations after cIV administration in NHL and B-ALL patients were simulated and compared to reported Css values. The observed vs predicted values can be seen in Tables 1 and 2.

**Table 1.**
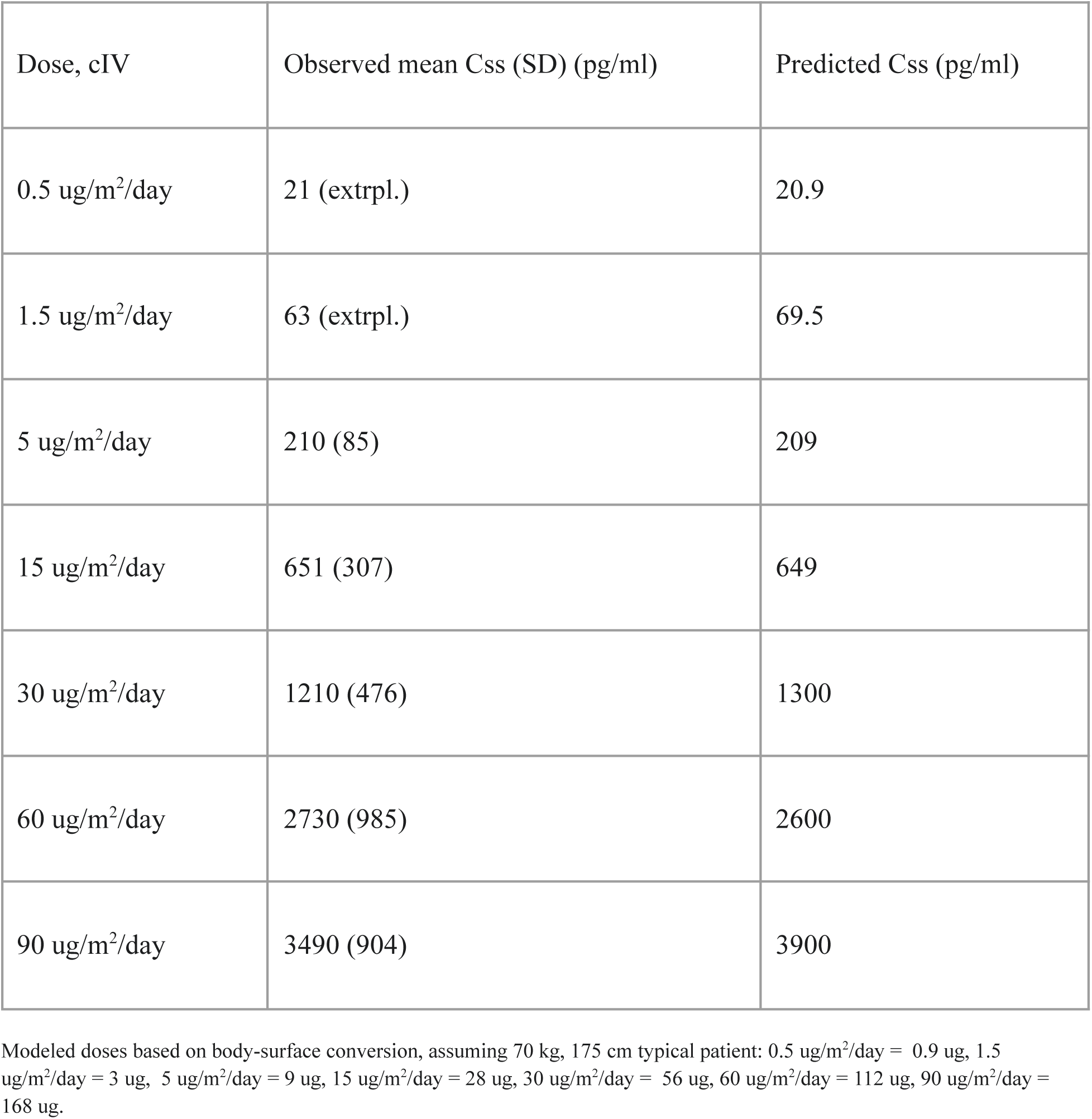
Css projections for cIV administration in NHL vs observed data. ^5^**. Extrpl. = not reported but extrapolated from the 5 ug/m^2^/day dose using linear dose-proportional extrapolation.**

| Dose, cIV | Observed mean C<sub>ss</sub> (SD) (pg/ml) | Predicted C<sub>ss</sub> (pg/ml) |
| --- | --- | --- |
| 0.5 ug/m<sup>2</sup>/day | 21 (extrpl.) | 20.9 |
| 1.5 ug/m<sup>2</sup>/day | 63 (extrpl.) | 69.5 |
| 5 ug/m<sup>2</sup>/day | 210 (85) | 209 |
| 15 ug/m<sup>2</sup>/day | 651 (307) | 649 |
| 30 ug/m<sup>2</sup>/day | 1210 (476) | 1300 |
| 60 ug/m<sup>2</sup>/day | 2730 (985) | 2600 |
| 90 ug/m<sup>2</sup>/day | 3490 (904) | 3900 |

Modeled doses based on body-surface conversion, assuming 70 kg, 175 cm typical patient: 0.5 ug/m<sup>2</sup>/day = 0.9 ug, 1.5 ug/m<sup>2</sup>/day = 3 ug, 5 ug/m<sup>2</sup>/day = 9 ug, 15 ug/m<sup>2</sup>/day = 28 ug, 30 ug/m<sup>2</sup>/day = 56 ug, 60 ug/m<sup>2</sup>/day = 112 ug, 90 ug/m<sup>2</sup>/day = 168 ug.

**Table 2.** Css projections for cIV administration in B-ALL vs observed data. Different values are from different studies, as indicated by the references.

| Dose, cIV, ug/day | Observed mean Css (SD), pg/ml | Predicted Css, pg/ml |
| --- | --- | --- |
| 9 | 228 (356)<sup>21</sup><br>229 (360)<sup>22</sup> | 204201 |
| 28 | 616 (537)<sup>21</sup><br>755 (455.5)<sup>21</sup><br>615 (538)<sup>22</sup> | 63527 |

Clinical trials with SC dosing in B-ALL were also conducted and the model simulations of PK were compared to the observed concentrations. Simulations of the approved cIV doses of 9 ug/day and 28 ug/day were also performed for comparison. The results can be seen in Figure 3.

**Figure 3.**
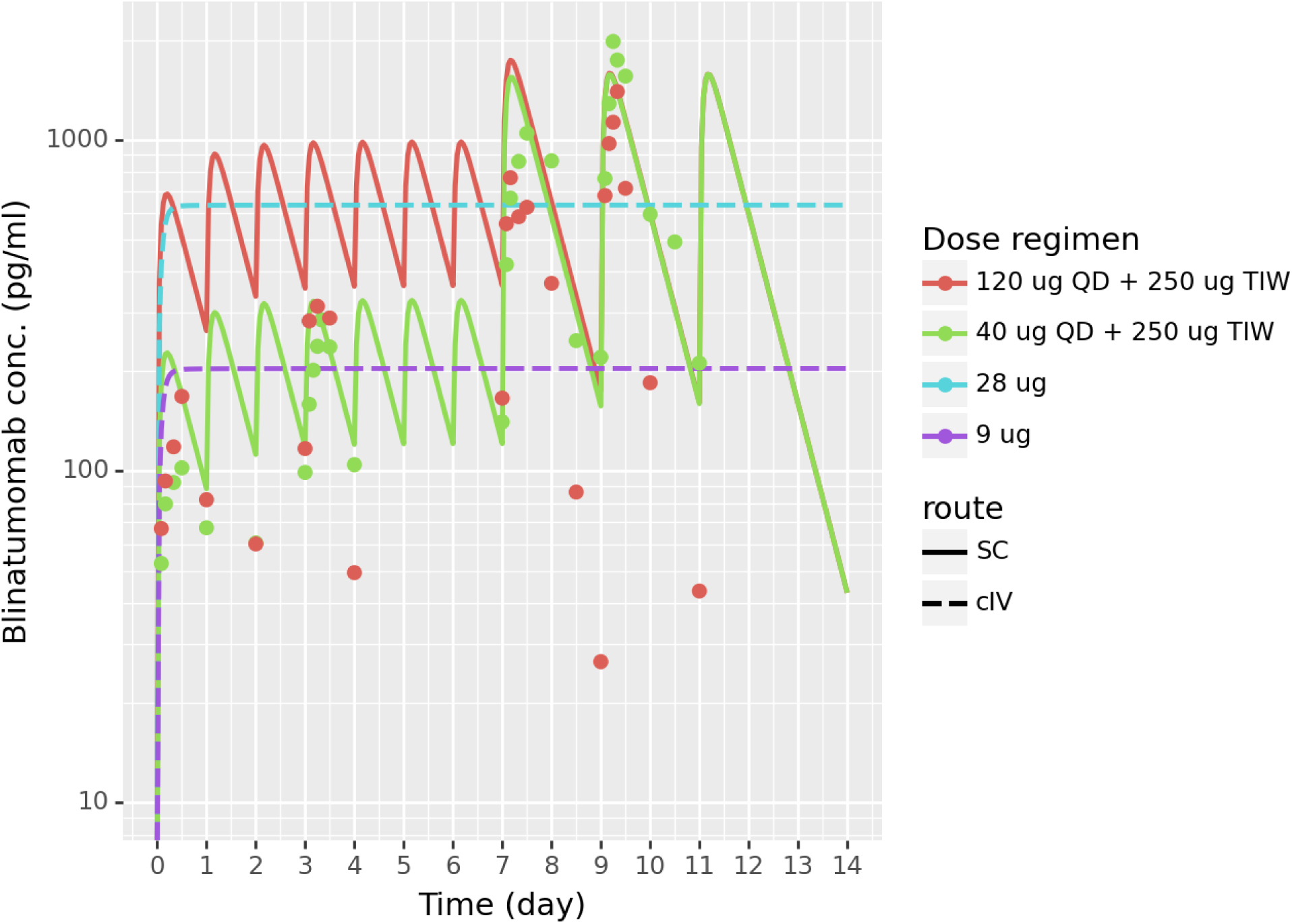
Blinatumomab model simulations vs observed B-ALL SC PK data ^6^. The model captures the 40 ug QD followed by 250 ug data well. The PK profile of the cohort with 120 ug QD starting dose could not be captured, since the reported responses were not dose-proportional to the lower dose cohort data. The 9 ug cIV and 28 ug cIV profiles were simulated for illustrative and comparison purposes.

With general agreement between projected and observed PK, simulations were run to compare the average trimer per T cell (TpT) formed between T cells and cancer cells at the site of action using the tested clinical doses. For NHL, the results can be seen in Figure 4 and for B-ALL in Figure 5. Different doses are in different colors, solid lines indicate SC administration, while dashed lines indicate cIV administration. Overall, predicted TpT at the clinical SC doses align with the predicted TpT at the higher infusion doses. Table 3 and Table 4 contain comparisons between average TpT at the site of action for administered doses for NHL and B-ALL, respectively.

**Figure 4.**
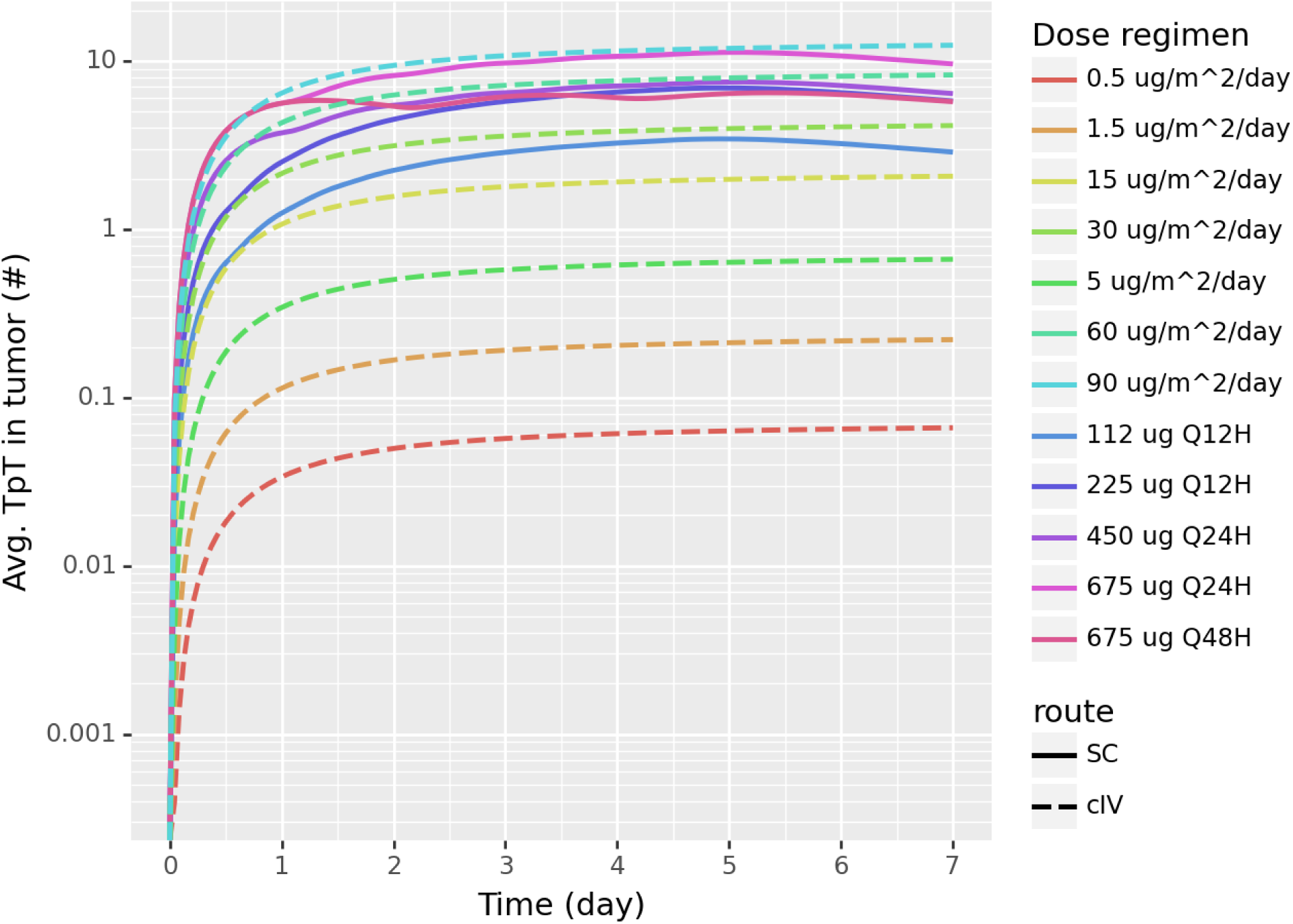
Blinatumomab NHL average TpT projections in the tumor with cIV and SC dosing regimens. The values shown are running averages - TpT AUC divided by the time point.

**Figure 5.**
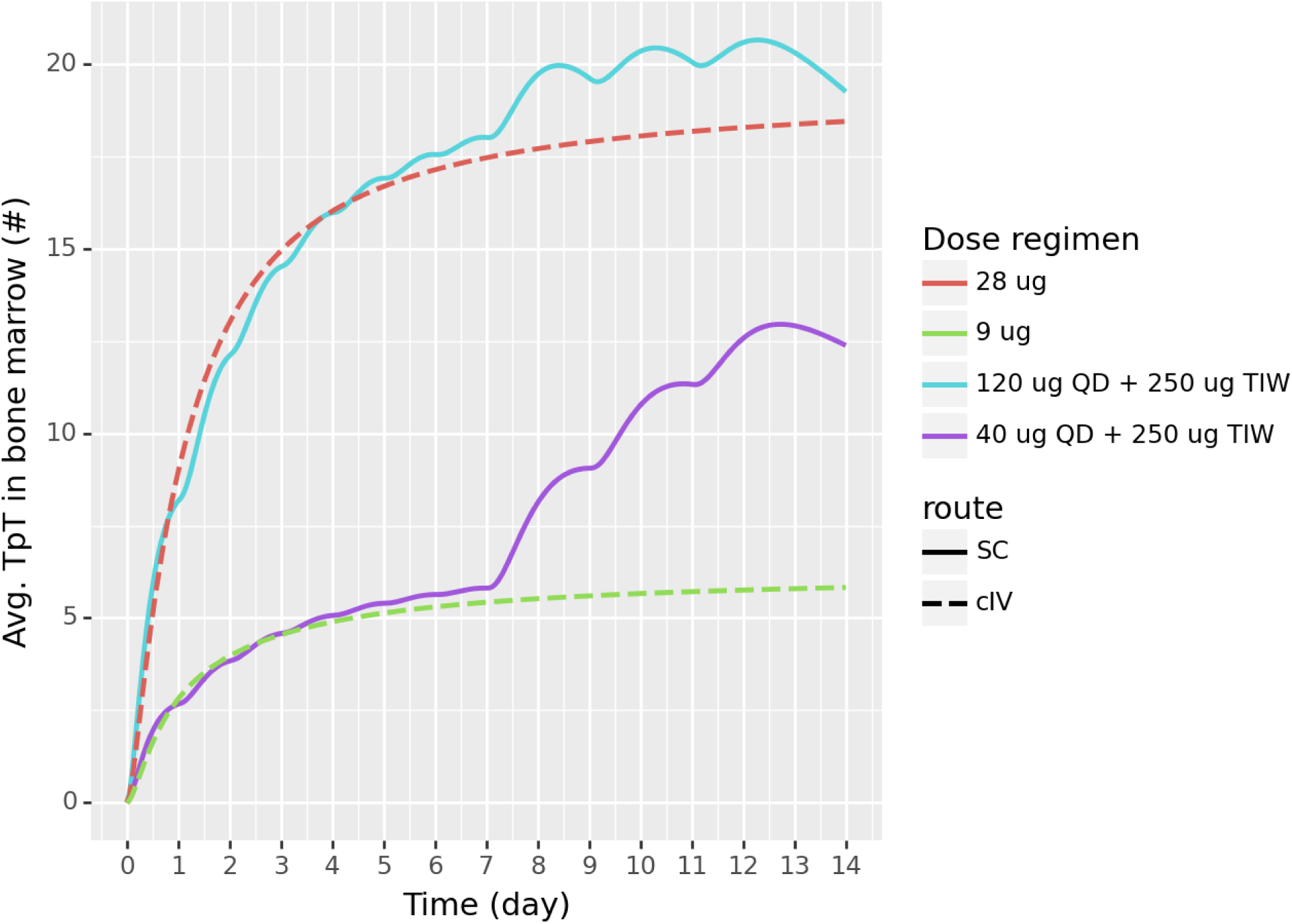
Blinatumomab B-ALL average TpT projections in the tumor with cIV and SC dosing regimens. The values shown running averages - TpT AUC divided by the time point.

**Table 3.** Avg TpT projection in NHL with cIV at day 28 and SC at day 7.

| Dose | Administration | Avg TpT in tumor, # |
| --- | --- | --- |
| 0.5 ug/m<sup>2</sup>/day | cIV | 0.071 |
| 1.5 ug/m<sup>2</sup>/day | cIV | 0.24 |
| 5 ug/m<sup>2</sup>/day | cIV | 0.71 |
| 15 ug/m<sup>2</sup>/day | cIV | 2.2 |
| 30 ug/m<sup>2</sup>/day | cIV | 4.4 |
| 60 ug/m<sup>2</sup>/day | cIV | 8.9 |
| 90 ug/m<sup>2</sup>/day | cIV | 13 |
| 112 ug Q12H | SC | 2.9 |
| 225 ug Q12H | SC | 5.8 |
| 450 ug QD | SC | 6.4 |
| 675 ug QD | SC | 9.6 |
| 675 ug Q2D | SC | 5.7 |

**Table 4.** Avg TpT projection with cIV and SC in B-ALL at day 14.

| Dose | Administration | Avg TpT, bone marrow<br>@ day 14 |
| --- | --- | --- |
| 9 ug/day | cIV | 5.83 |
| 28 ug/day | cIV | 18.5 |
| 40 ug QD + 250 ug TIW | SC | 12.4 |
| 120 ug QD + 250 ug TIW | SC | 19.3 |

To compare the TpT for cIV regimens against potential SC regimens, a range of SC administered doses were simulated and the results at day 28, the end of one typical cycle of blinatumomab, were compared to the day 28 steady state results after cIV administration. For B-ALL, 28 ug/day cIV was predicted to achieve 18.9 average TpT by day 28. With SC administration, doses of 115 ug QD and 225 ug Q2D were projected to achieve the same amount of trimers (See Table 5).

**Table 5.** Avg TpT projection with SC QD or Q2D dosing for 28 days vs equivalent cIV doses in NHL and B-ALL.

| Indication | Dose (SC) | Avg TpT at the site of action by day 28 achieved by SC dose, # | Equivalent dose, cIV | Avg TpT at the site of action by day 28 achieved by cIV dose, # |
| --- | --- | --- | --- | --- |
| NHL | 450 ug QD | 8.85 | 112 ug/day | 8.90 |
|  | 890 ug Q2D | 8.89 |  |  |
| B-ALL | 115 ug QD | 19.3 | 28 ug/day | 18.9 |
|  | 225 ug Q2D | 19.1 |  |  |

Comparison of the number of trimers achieved with the three doses can be seen in Figure 6. For NHL, the dose of 60 ug/m^2^/day cIV, equivalent to 112 ug/day cIV as administered in subsequent studies, was chosen as the dose for which to find an equivalent dose with SC administration. 112 ug/day is projected to achieve 8.9 TpT in the tumor and the same amount of trimers are achieved with 450 ug QD and 890 ug Q2D. Comparison of the number of trimers achieved with the three doses can be seen in Figure 7. Table 5 contains the projected average site-of-action number of trimers for each dose and indication.

**Figure 6.**
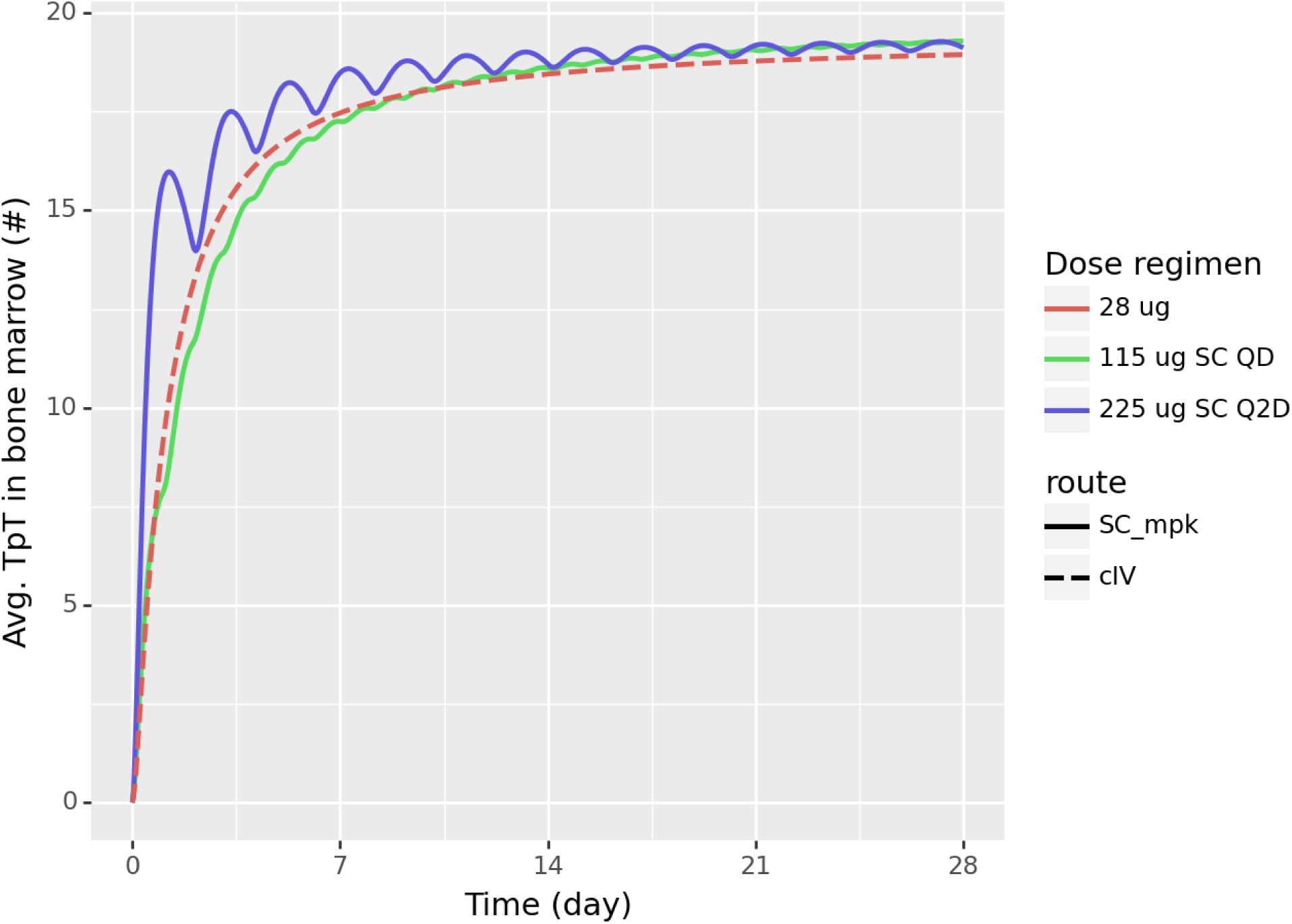
Blinatumomab simulated Avg TpT in the bone marrow for predicted effective dose with QD or Q2D SC dosing for 28 days in B-ALL. The doses are picked so that the Avg TpT matches 28 ug/day cIV dose.

**Figure 7.**
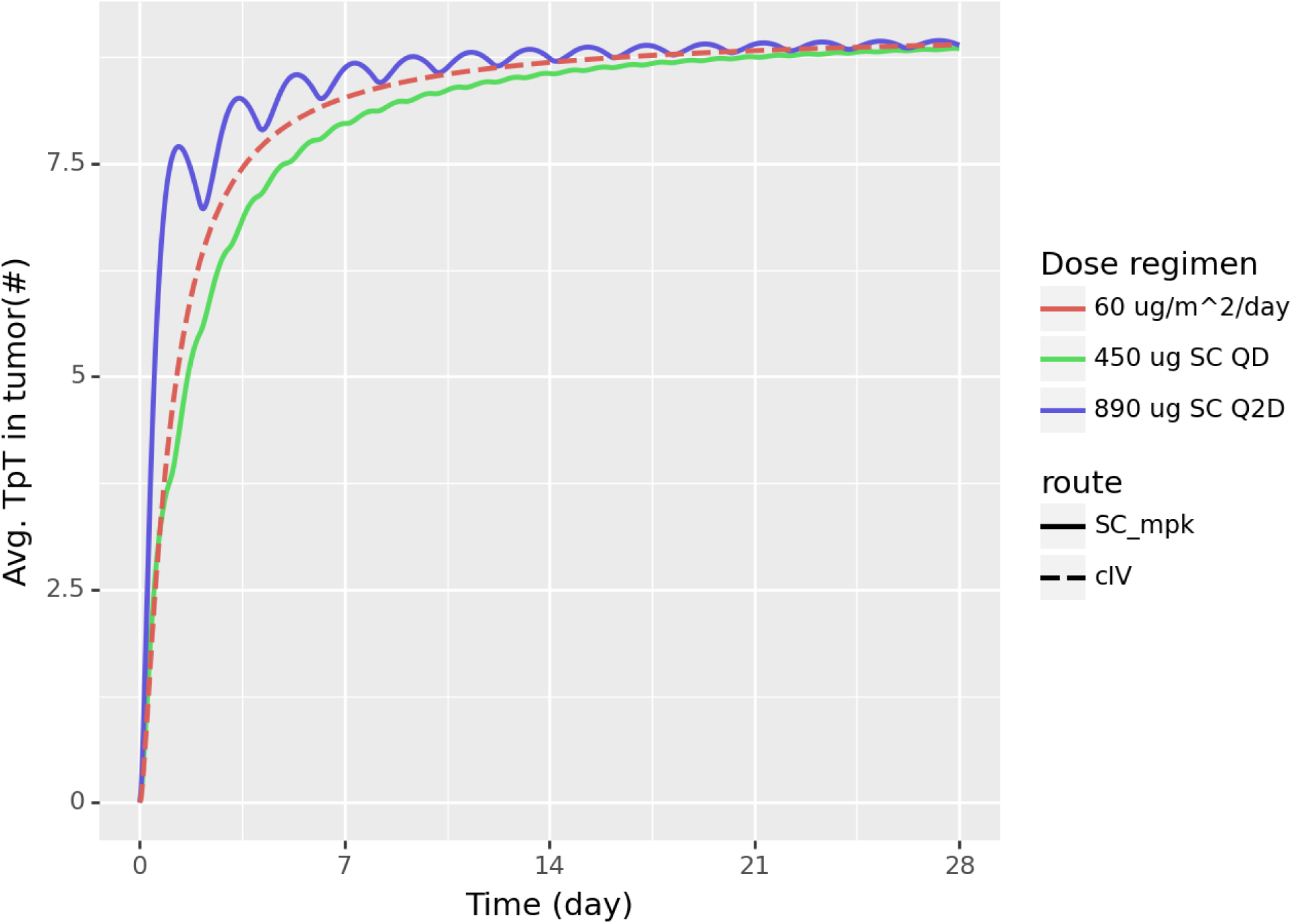
Blinatumomab simulated Avg TpT in the tumor for predicted effective dose with QD or Q2D SC dosing for 28 days in NHL. The doses are picked so that the Avg TpT matches 112 ug/day cIV dose.

A one-at-a-time sensitivity analysis was performed to explore the effect of fold-change of each model parameter on the average TpT at the site of action at day 28. For each indication, two fixed doses were explored - the cIV dose used to evaluate SC equivalence (112 ug/day for NHL and 28 ug/day for B-ALL) and the equivalent SC QD doses from the result above (450 ug for NHL and 115 ug for B-ALL). The fold-change for each parameter varied between 2, 3, and 10, depending on the parameter type and the level of uncertainty in parameter estimation. For example, the central volume was only varied two-fold in either direction, since the plasma volume is not expected to vary substantially for the typical individual. In contrast, the cross-linking on-rate constant was varied ten-fold since the value has not been experimentally determined and in the case that it had been measured, variations between in vitro and in vivo measurements could be expected (hence why the affinities are also varied by ten-fold). Figures <u>S2</u>-<u>S3</u> and <u>S4</u>-<u>S5</u> present Tornado plots of the results for NHL and B-ALL, respectively. The parameters were ranked based on the absolute difference in average TpT resulting from simulations with the top and the bottom values of the parameter. Only the top 15 most sensitive parameters are pictured. The most sensitive parameters are different for the two indications. For B-ALL, the most sensitive parameters are the on-rate constants, the affinity to CD3, the number of CD3 molecules per T cell and the elimination half-life. For NHL, the most sensitive parameters are the number of T cells, the distribution of blinatumomab to the tumor, the tumor volume, and the internalization rate of CD19.

When comparing the results between the cIV and the SC doses (Figures <u>S2</u> vs <u>S3</u> for NHL and <u>S4</u> vs <u>S5</u> for B-ALL), besides bioavailability, which has no effect on cIV simulations, the order of the sensitive parameters and the magnitude by which they change the TpT projections is almost identical within each indication. This indicates that even if parameters change, a similar SC dose to what is currently projected will achieve the equivalent TpT as its cIV counterpart, making the current dose-equivalence results insensitive to parameter fluctuations within the folds explored.

## Discussion

The TCE model constructed here was used to bridge the currently used cIV and the explored SC dosing regimens for blinatumomab by exploring the effect of each on a model-based metric of effect - average trimers per T cell at the site of action. The two SC dosing regimens that were used were QD and Q2D. Based on the modeling results, for B-ALL, in order to achieve the same TpT as 28 ug/day cIV, 115 ug SC QD or 225 ug SC Q2D would be required. For NHL, in order to achieve the same TpT as 112 ug/day cIV, 450 ug SC QD or 890 ug SC Q2D would be required. These clinical doses align well with what has already been explored in the clinic, which validates the modeling results. For B-ALL, 120 ug QD, followed by 250 ug TIW has been explored in a clinical study ^6^. For NHL, a 450 ug SC QD regimen was also a part of a clinical study ^7^. Furthermore, the equivalent SC dose projections are insensitive to parameter fluctuations as explored by sensitivity analysis, which increases the robustness of the dose projections.

The model does not address the full complexity of either disease and the full effect of drug intervention on patients. Cellular dynamics, beyond B-cell depletion, which was incorporated for capturing the TMDD dynamics more accurately, have not been explored. T-cell activation, proliferation, and site-of-action infiltration, along with cancer cell death have not been explored - they could be an important component of both effective dose predictions and bridging the cIV to SC dosing. The effect of blinatumomab on cytokine release syndrome (CRS) and other adverse events was also not modeled, while those considerations are crucial when making decisions on changes in administration. Furthermore, in the clinic, in order to mitigate the effect of CRS on patients, a step-wise dosing approach is utilized - the patients are not dosed at the final efficacious dose but a lower dose is initially used to “prime” the immune system ^23,24^. For example, in B-ALL, the first month-long cycle with blinatumomab is at 9 ug/day cIV, which is then increased to 28 ug/day in subsequent cycles ^1^. However, to our knowledge, comprehensive dynamics of CRS and proliferation data are not available in the literature for blinatumomab and particularly for the SC dosing regimens. Hence, while the model can easily be expanded to incorporate these dynamics, they are beyond the scope of the current study.

It is unclear whether average trimer per T cell is the correct metric to explore for an effective dose, considering the speed of B-cell depletion, the process of activating T cells and subsequent death of target cells appear to be relatively quick when compared to the length of treatment cycle (a few hours vs a month). Therefore, a maximum trimer metric may be a reasonable approach, which would lead to projecting lower effective doses with SC administration when compared to cIV doses (not shown here). However, given cancer cell resilience and variable patient responses, a trough trimer metric is not out of the question, which would lead to higher projected effective doses with SC administration. More blinatumomab clinical data with SC dosing is needed to guide the appropriate metric for determining safe and effective doses.

The sensitivity analysis results for the model projections for B-ALL and NHL revealed that when access to the site of action is limited, as is the case with NHL in the model, overcoming this limitation and increasing distribution is more important than the act of forming trimers. However, when the site of action provides easy access, as is the case with bone marrow in B-ALL, the actual binding and forming trimers is more sensitive.

Furthermore, the model predicts fewer trimers being formed in the NHL site of action than B-ALL. This prediction aligns well with the higher blinatumomab doses explored in NHL vs B-ALL in the clinic. While distribution is likely not the only factor that makes NHL more challenging for blinatumomab than B-ALL, the model assumption that the drug concentration in the tumor is lower than that in the bone marrow and hence leads to fewer trimers formed in the tumor versus the bone marrow, aligns with clinical observations.

Further work exploring the model results and comparing safety profiles between cIV and SC dosing is necessary for a robust understanding for optimal dosing regimens. Those explorations and predictions can be achieved with the current model but more data are needed to validate the model predictions.

## Study Highlights

Articles should include a Study Highlights section after the Discussion in the manuscript text. The highlights section should include and answer each of the questions below. The entire section, not including the questions, should be under 150 words.

- What is the current knowledge on the topic? To our knowledge no external publication attempt has been made to bridge the cIV to SC dosing for blinatumomab, hence the method for bridging has not been explored in the literature.
- What question did this study address? The study showcases a model that predicts the doses with SC administration that can achieve the same formation of CD3:blinatumomab:CD19 molecules, hypothesized to be directly related to efficacy, as the currently approved cIV administration for blinatumomab.
- What does this study add to our knowledge? The study suggests which SC doses should be explored in the clinic for blinatumomab. Given that the SC administration is novel for this molecule, the model’s predictions can be applied to future clinical studies.
- How might this change clinical pharmacology or translational science? T-cell engagers are challenging molecules to model and predict safe and efficacious doses in the clinic. The study presents a potential method for bridging IV to SC dosing regimens for TCEs.

## Data Availability

All data produced in the present study are available upon reasonable request to the authors

## Acknowledgements

The authors would like to thank Victor Chang for his contribution to the study of T-cell engagers at Applied BioMath.

## Author Contributions

GK, SH, JA, DF, and JG wrote the manuscript; GK and JG designed the research; GK, SH, and DF performed the research; GK and SH analyzed the data.

## Supplementary Information

**Table S1:**
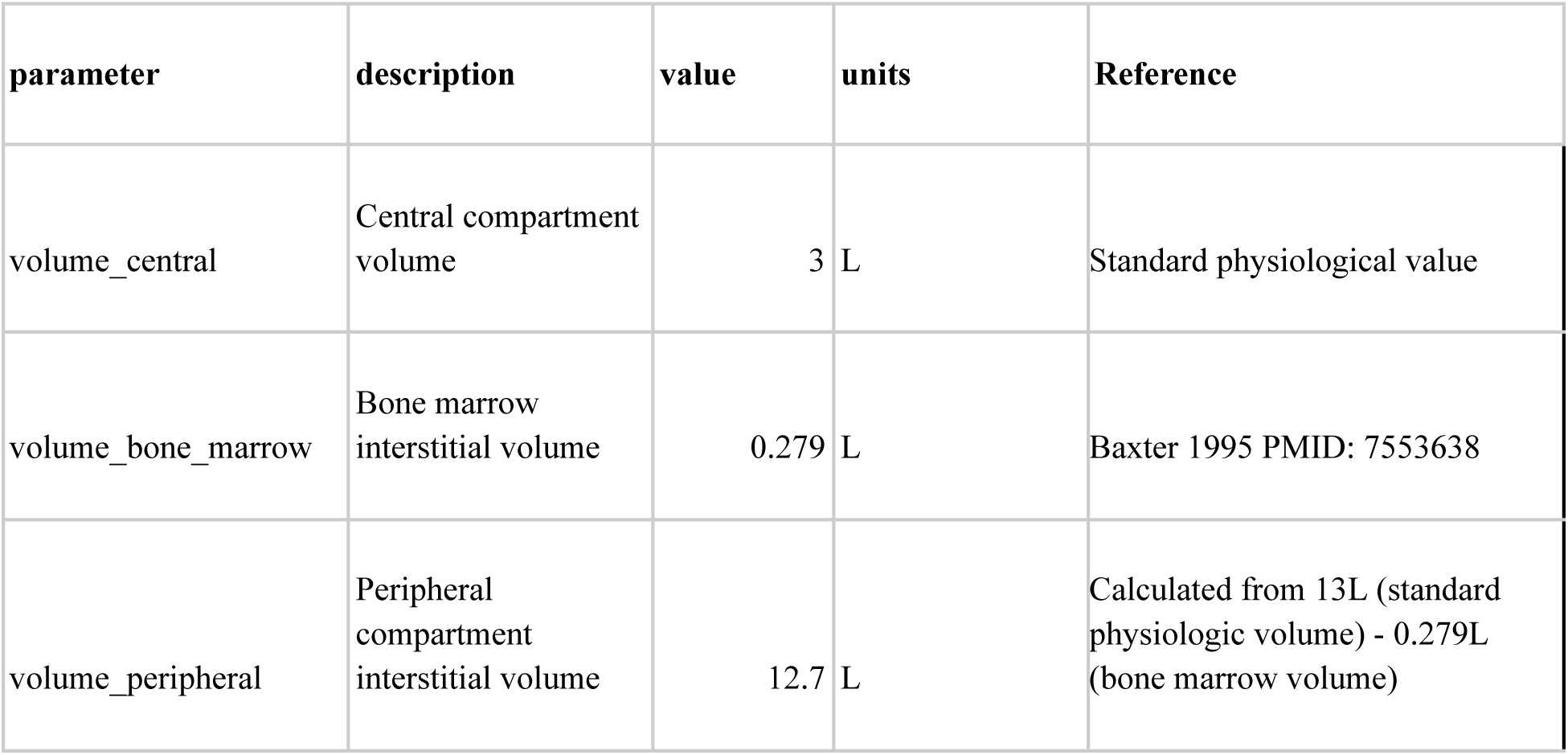
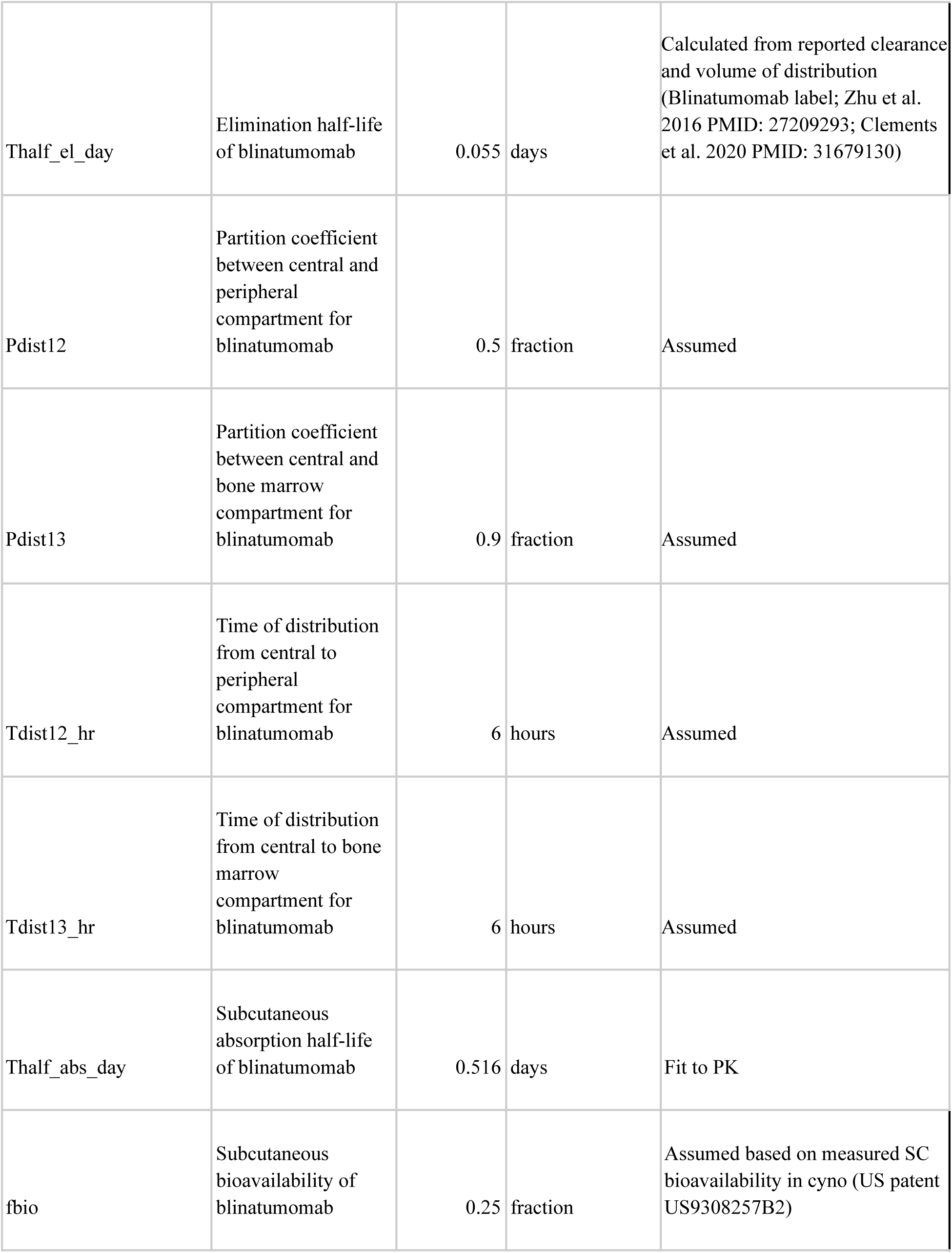
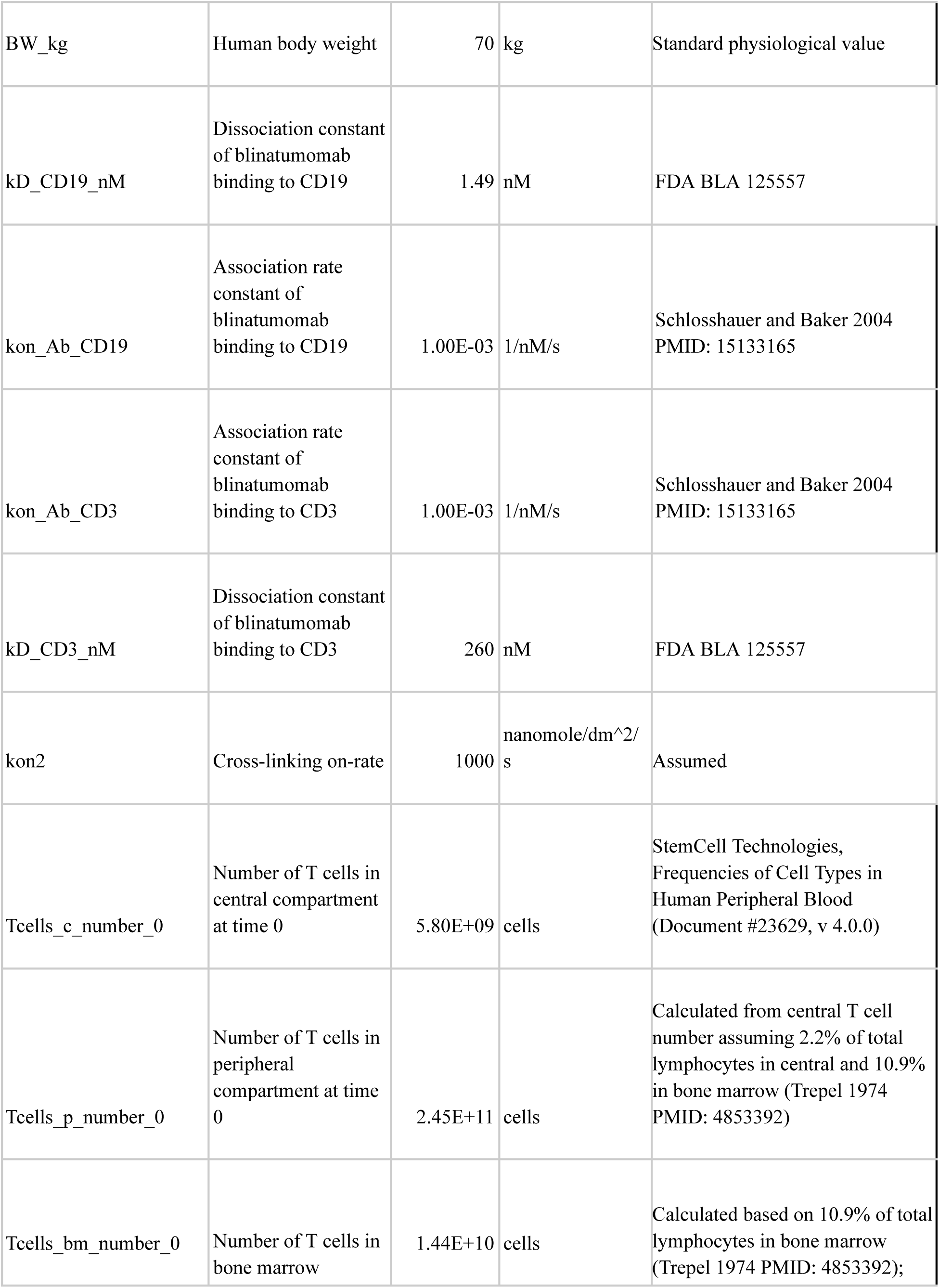
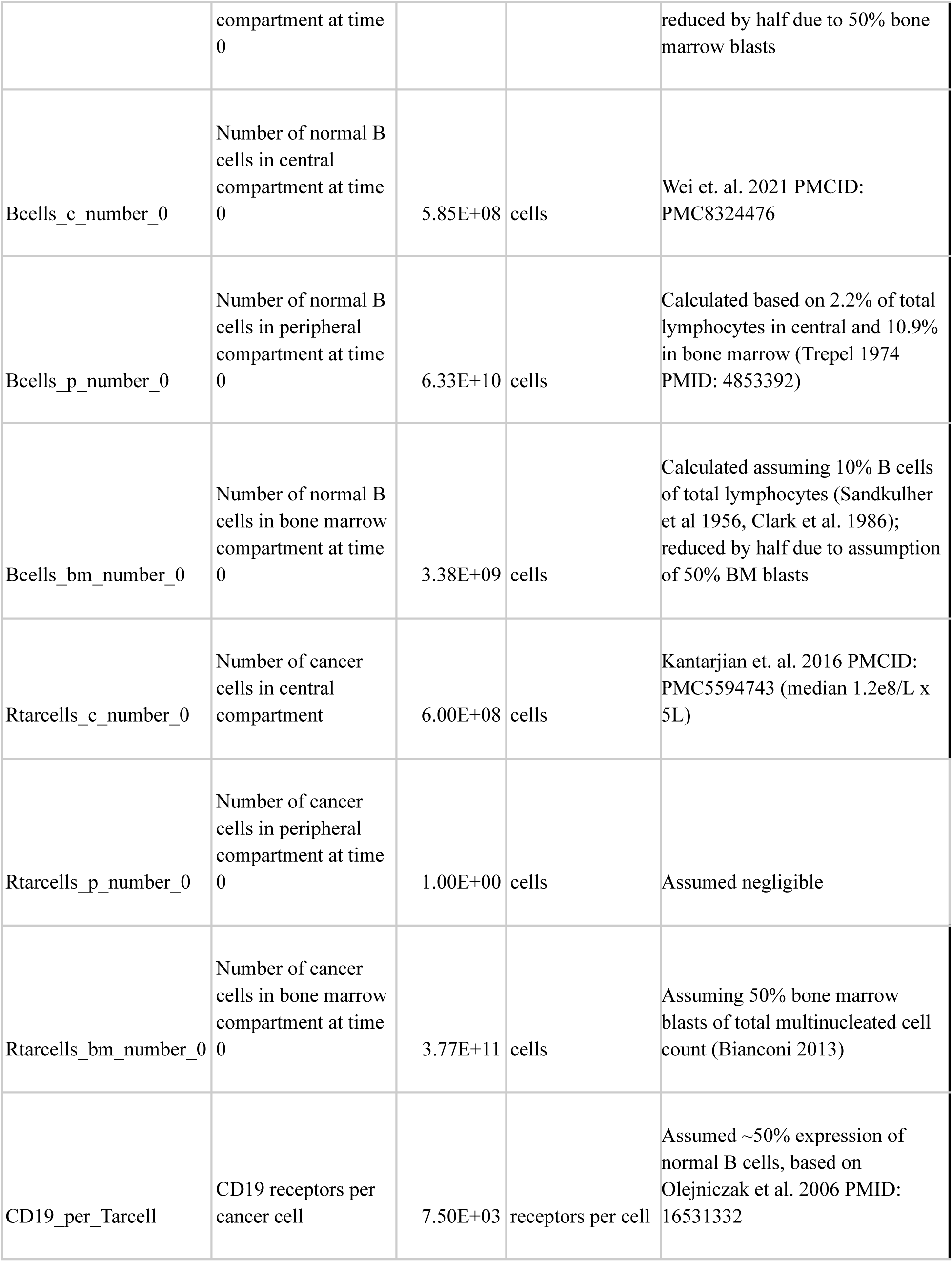
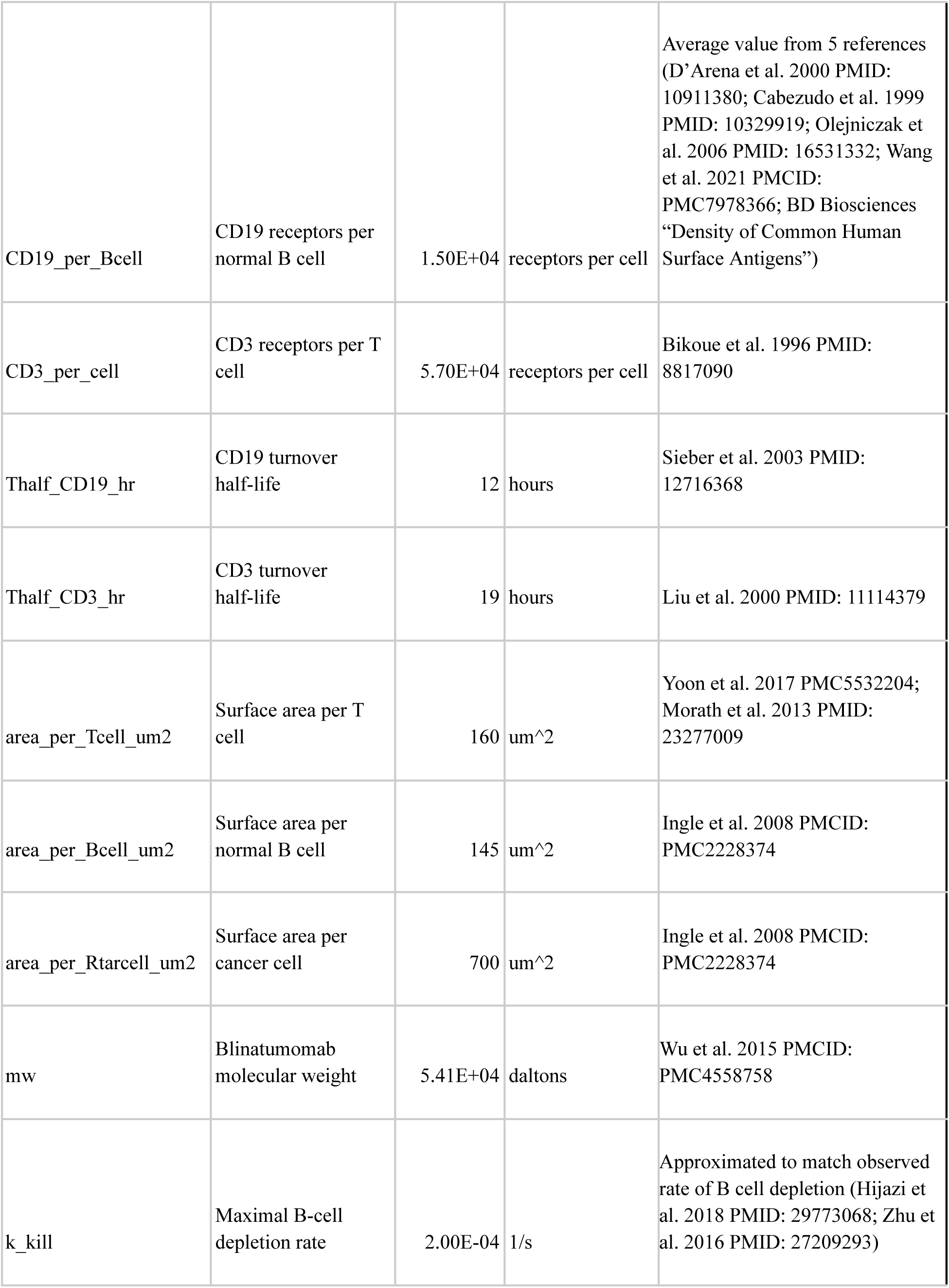
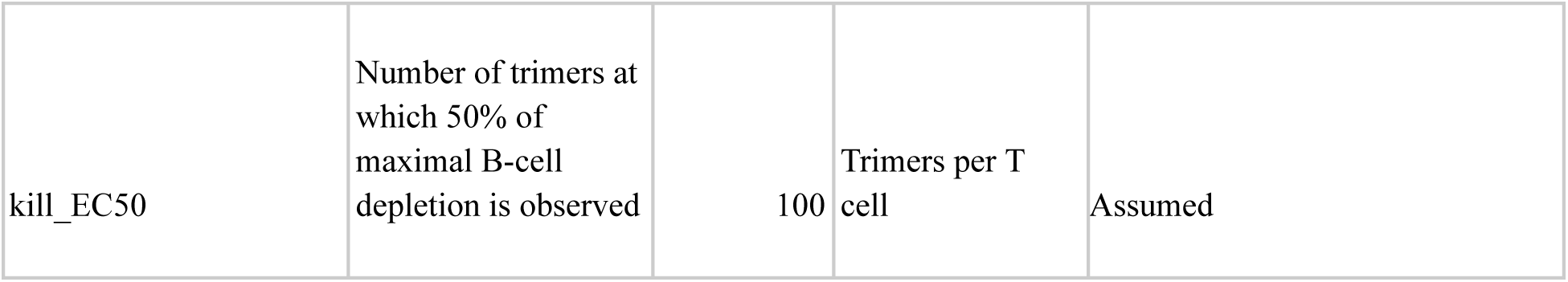
B-ALL Model parameters.

**Table S2:** NHL Model parameters.

| parameter | description | value | units | Reference |
| --- | --- | --- | --- | --- |
| volume_central | Central compartment volume | 3 | L | Standard physiological value |
| volume_tumor | Tumor interstitial volume | 0.012 | L | Calculated based on assumed ~5 cm tumor diameter (~60 mL volume) and 20% interstitial fraction |
| volume_peripheral | Peripheral compartment interstitial volume | 13 | L | Standard physiological value |
| Thalf_el_day | Elimination half-life of blinatumomab | 0.055 | days | Calculated from reported clearance and volume of distribution (Blinatumomab label; Zhu et al. 2016 PMID: 27209293; Clements et al. 2020 PMID: 31679130) |
| Pdist12 | Partition coefficient between central and peripheral compartment for blinatumomab | 0.5 | fraction | Assumed |
| Pdist13 | Partition coefficient between central and tumor compartment for blinatumomab | 0.3 | fraction | Assumed |
| Tdist12_hr | Distribution half-life from central to peripheral compartment for blinatumomab | 6 | hours | Assumed |
| Tdist13_hr | Distribution half-life from central to tumor compartment for blinatumomab | 34 | hours | Assumed |
| Thalf_abs_day | Subcutaneous absorption half-life of blinatumomab | 0.516 | days | Fit to PK |
| fbio | Subcutaneous bioavailability of blinatumomab | 0.25 | fraction | Assumed to be the same as SC bioavailability in cyno (US patent US9308257B2) |
| BW_kg | Human body weight | 70 | kg | Standard physiological value |
| kD_CD19_nM | Dissociation constant of blinatumomab binding to CD19 | 1.49 | nM | FDA BLA 125557 |
| kon_Ab_CD19 | Association rate constant of blinatumomab binding to CD19 | 0.001 | 1/nM/s | Schlosshauer and Baker 2004<br>PMID: 15133165 |
| kon_Ab_CD3 | Association rate constant of blinatumomab binding to CD3 | 0.001 | 1/nM/s | Schlosshauer and Baker 2004<br>PMID: 15133165 |
| kD_CD3_nM | Dissociation constant of blinatumomab binding to CD3 | 260 | nM | FDA BLA 125557 |
| kon2 | Cross-linking on-rate | 1000 | nanomole/dm<sup>2</sup>/s | Assumed |
| Tcells_c_number_0 | Number of T cells in central compartment at time 0 | 5.8E+09 | cells | StemCell Technologies, Frequencies of Cell Types in Human Peripheral Blood (Document #23629, v 4.0.0) |
| Tcells_p_number_0 | Number of T cells in peripheral compartment at time 0 | 2.6E+11 | cells | Calculated based on 2.2% of total lymphocytes in central (Trepel 1974 PMID: 4853392) |
| Tcells_t_number_0 | Number of T cells in tumor compartment at time 0 | 1.84E+09 | cells | Calculated based on median 30.6% of tumor cells in follicular lymphoma (Wahlin et al. 2007 PMID: 17255259), 60 mL tumor volume and tumor cell density of 1E8 cells/mL (Del Monte 2009) |
| Bcells_c_number_0 | Number of normal B cells in central compartment at time 0 | 1.5E+09 | cells | StemCell Technologies, Frequencies of Cell Types in Human Peripheral Blood (Document #23629, v 4.0.0) |
| Bcells_p_number_0 | Number of normal B cells in peripheral compartment at time 0 | 6.67E+10 | cells | Calculated based on 2.2% of total lymphocytes in central (Trepel 1974 PMID: 4853392) |
| Bcells_t_number_0 | Number of normal B cells in tumor compartment at time 0 | 1 | cells | Assumed negligible |
| Rtarcells_c_number_0 | Number of cancer cells in central compartment | 1 | cells | Assumed negligible |
| Rtarcells_p_number_0 | Number of cancer cells in peripheral compartment at time 0 | 1 | cells | Assumed negligible |
| Rtarcells_t_number_0 | Number of cancer cells in tumor compartment at time 0 | 4.02E+09 | cells | Calculated based on median 67% of tumor cells in follicular lymphoma (Wahlin et al. 2007 PMID: 17255259), 60 mL tumor volume and tumor cell density of 1E8 cells/mL (Del Monte 2009) |
| CD19_per_Tarcell | CD19 receptors per cancer cell | 5,250 | receptors per cell | Assumed ~35% of normal B cells, based on measurements in follicular lymphoma, diffuse large B-cell lymphoma, and mantle cell lymphoma (D'Arena et al. 2000 PMID: 10911380; Cabezudo et al. 1999 PMID: 10329919; Olejniczak et al. 2006 PMID: 16531332) |
| CD19_per_Bcell | CD19 receptors per normal B cell | 15,000 | receptors per cell | Average value from 5 references (D'Arena et al. 2000 PMID: 10911380; Cabezudo et al. 1999 PMID: 10329919; Olejniczak et al. 2006 PMID: 16531332; Wang et al. 2021 PMCID: PMC7978366; BD Biosciences "Density of Common Human Surface Antigens") |
| CD3_per_cell | CD3 receptors per T cell | 57,000 | receptors per cell | Bikoue et al. 1996 PMID: 8817090 |
| Thalf_CD19_hr | CD19 turnover half-life | 12 | hours | Assumed, based on Sieber et al. 2003 PMID: 12716368 |
| Thalf_CD3_hr | CD3 turnover half-life | 19 | hours | Liu et al. 2000 PMID: 11114379 |
| area_per_Tcell_um2 | Surface area per T cell | 160 | um<sup>2</sup> | Yoon et al. 2017 PMC5532204; Morath et al. 2013 PMID: 23277009 |
| area_per_Bcell_um2 | Surface area per normal B cell | 145 | um<sup>2</sup> | Ingle et al. 2008 PMCID: PMC2228374 |
| area_per_Rtarcell_um2 | Surface area per cancer cell | 700 | um<sup>2</sup> | Ingle et al. 2008 PMCID: PMC2228374 |
| mw | Blinatumomab molecular weight | 54,100 | daltons | Wu et al. 2015 PMCID: PMC4558758 |
| k_kill | Maximal B-cell depletion rate | 0.0002 | 1/s | Approximated to match observed rate of B cell depletion (Hijazi et al. 2018 PMID: 29773068; Zhu et al. 2016 PMID: 27209293) |
| kill_EC50 | Number of trimers at which 50% of maximal B-cell depletion is observed | 100 | Trimers per T cell | Assumed |

**Figure S1.**
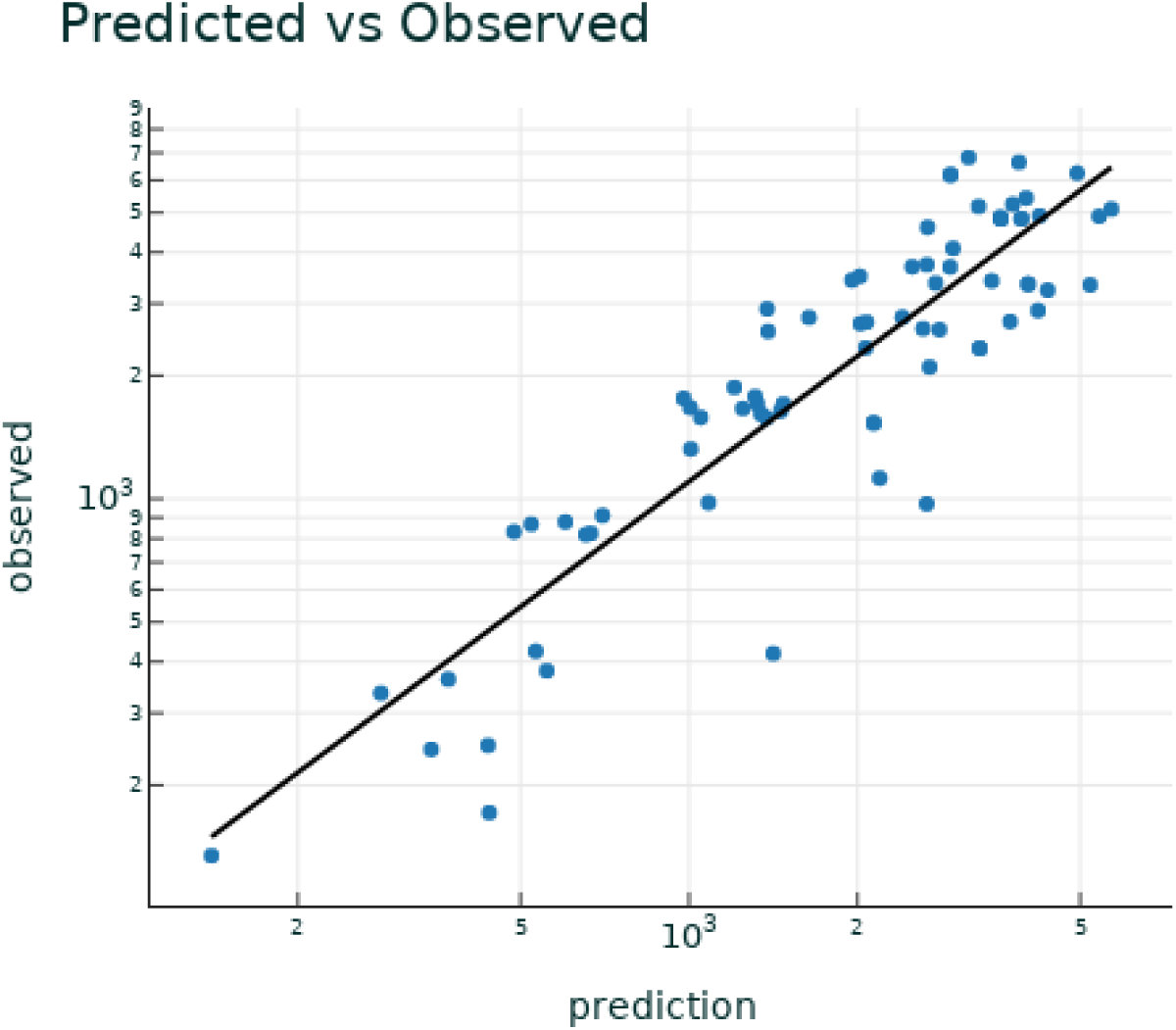
**Observed vs Predicted plot for the calibration of the model to NHL PK data.**

**Figure S2.**
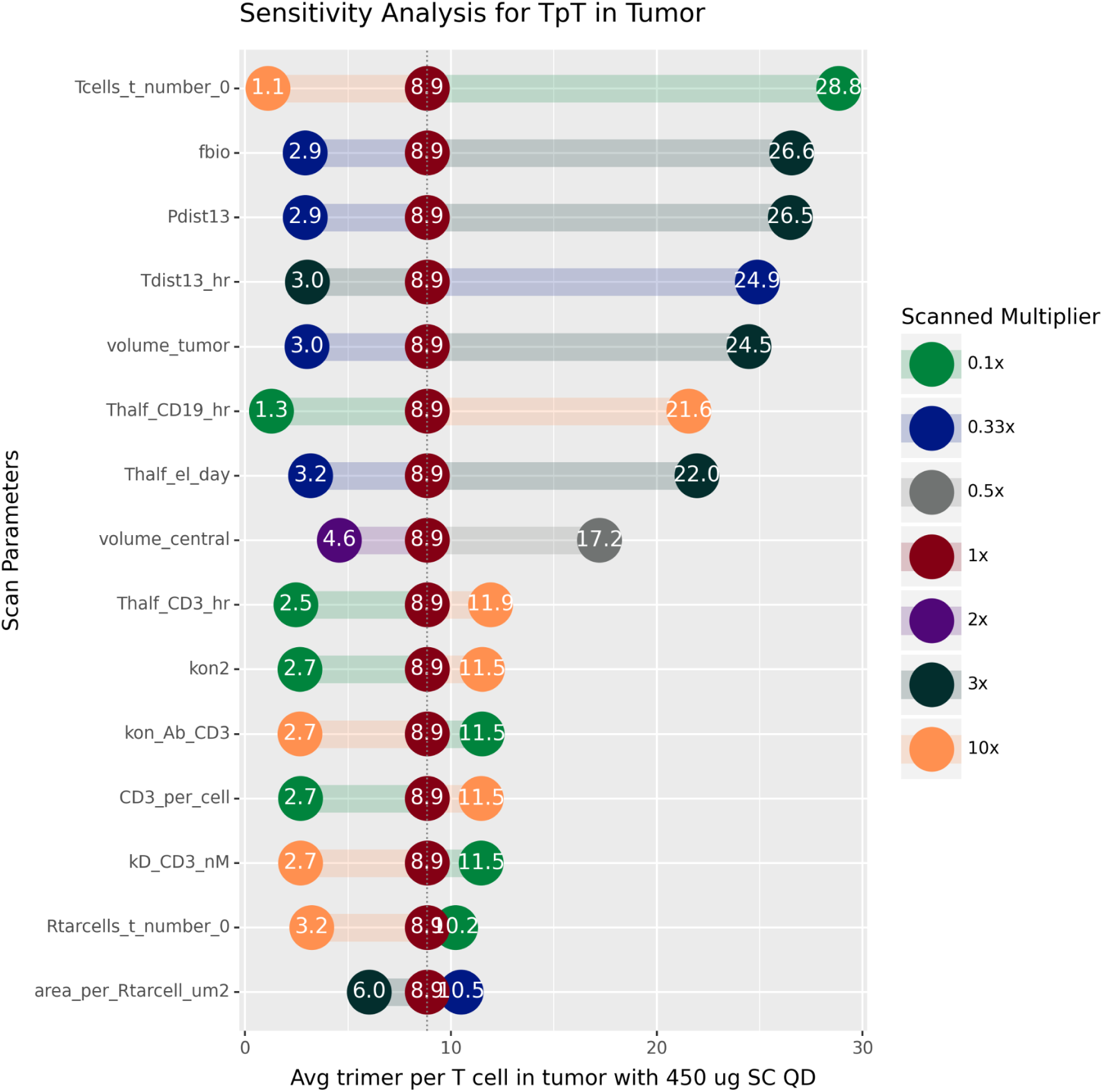
Results from sensitivity analysis with SC dosing in NHL patients. The dosing schedule was fixed at 450 ug SC QD. The parameters were varied one at a time by several fold, as indicated in the legend, and the average TpT at day 28 was reported. The parameters are in rank order based on the highest absolute difference in the results between the top and bottom parameter value. Only the top fifteen values are shown. A description of the parameters and their nominal values are in <u>Table S2</u>.

**Figure S3.**
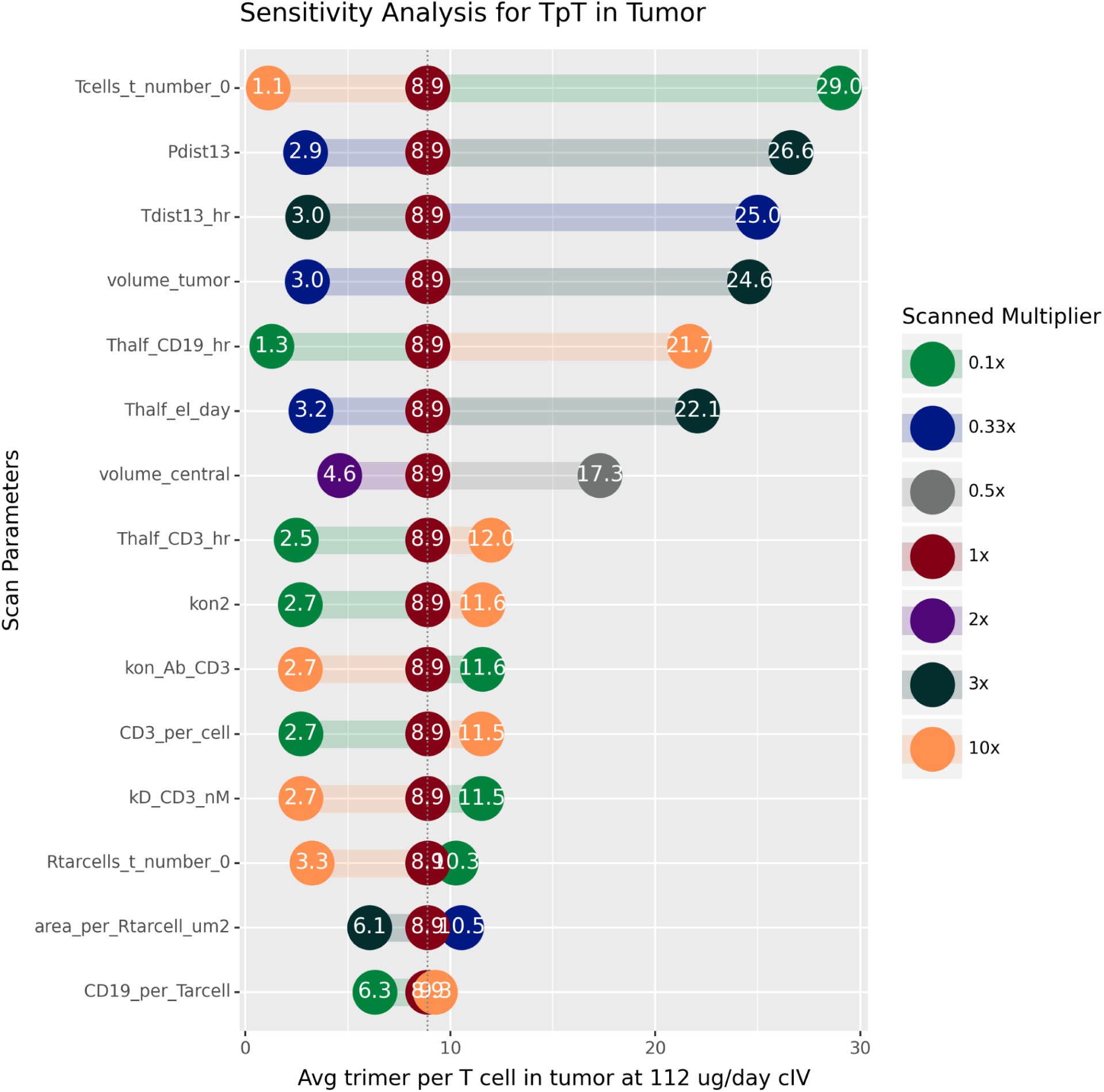
Results from sensitivity analysis with cIV dosing in NHL. The dosing schedule was fixed at 112 ug/day. The parameters were varied one at a time by several fold, as indicated in the legend, and the average TpT at day 28 was recorded. The parameters are in rank order based on the highest absolute difference in the results between the top and bottom parameter value. Only the top fifteen values are shown. A description of the parameters and their nominal values are in <u>Table S2</u>.

**Figure S4.**
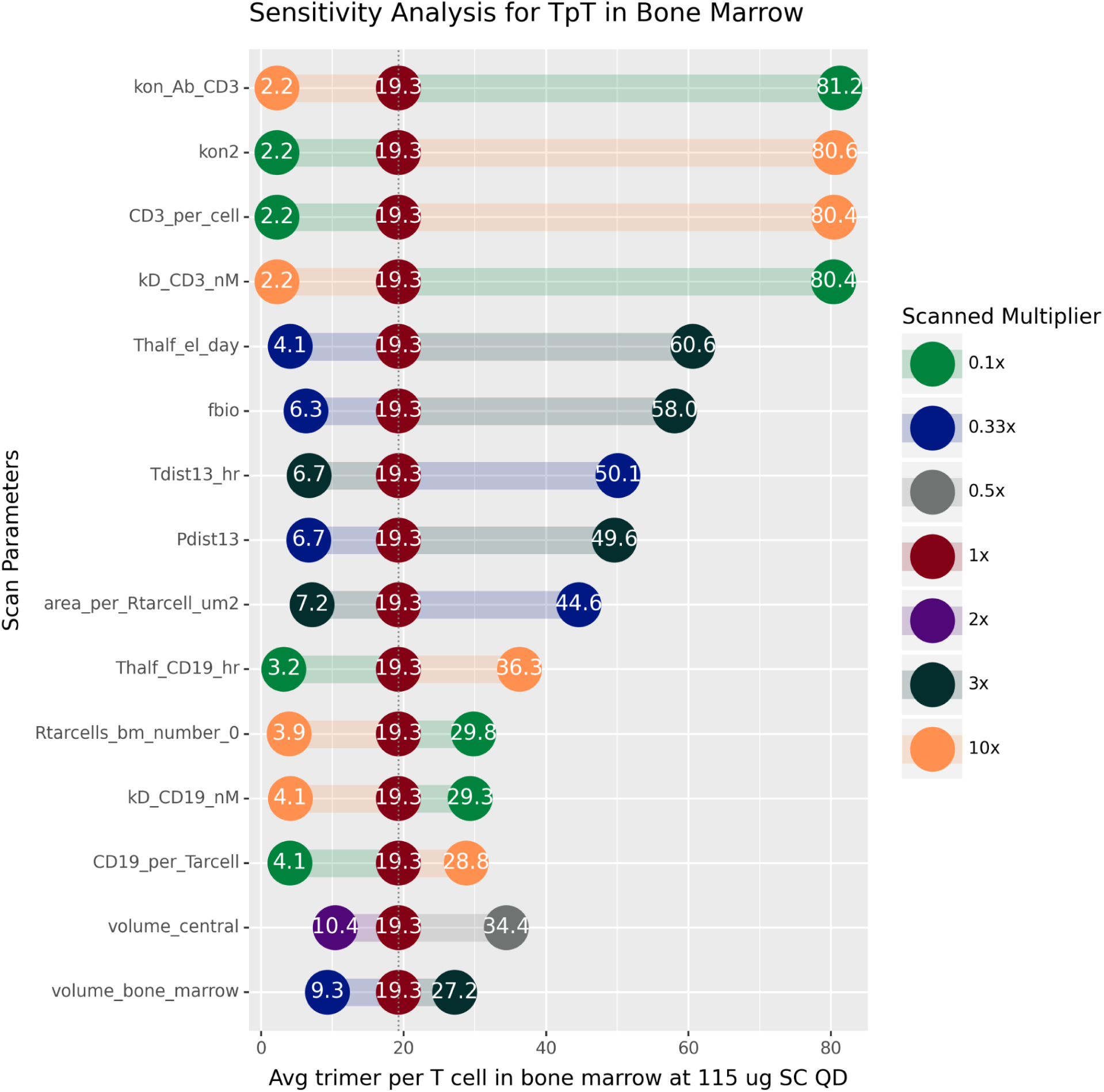
Results from sensitivity analysis with SC dosing in B-ALL. The dosing schedule was fixed at 115 ug SC QD. The parameters were varied one at a time by several fold, as indicated in the legend, and the average TpT at day 28 was recorded. The parameters are in rank order based on the highest absolute difference in the results between the top and bottom parameter value. Only the top fifteen values are shown. A description of the parameters and their nominal values are in <u>Table S1</u>.

**Figure S5.**
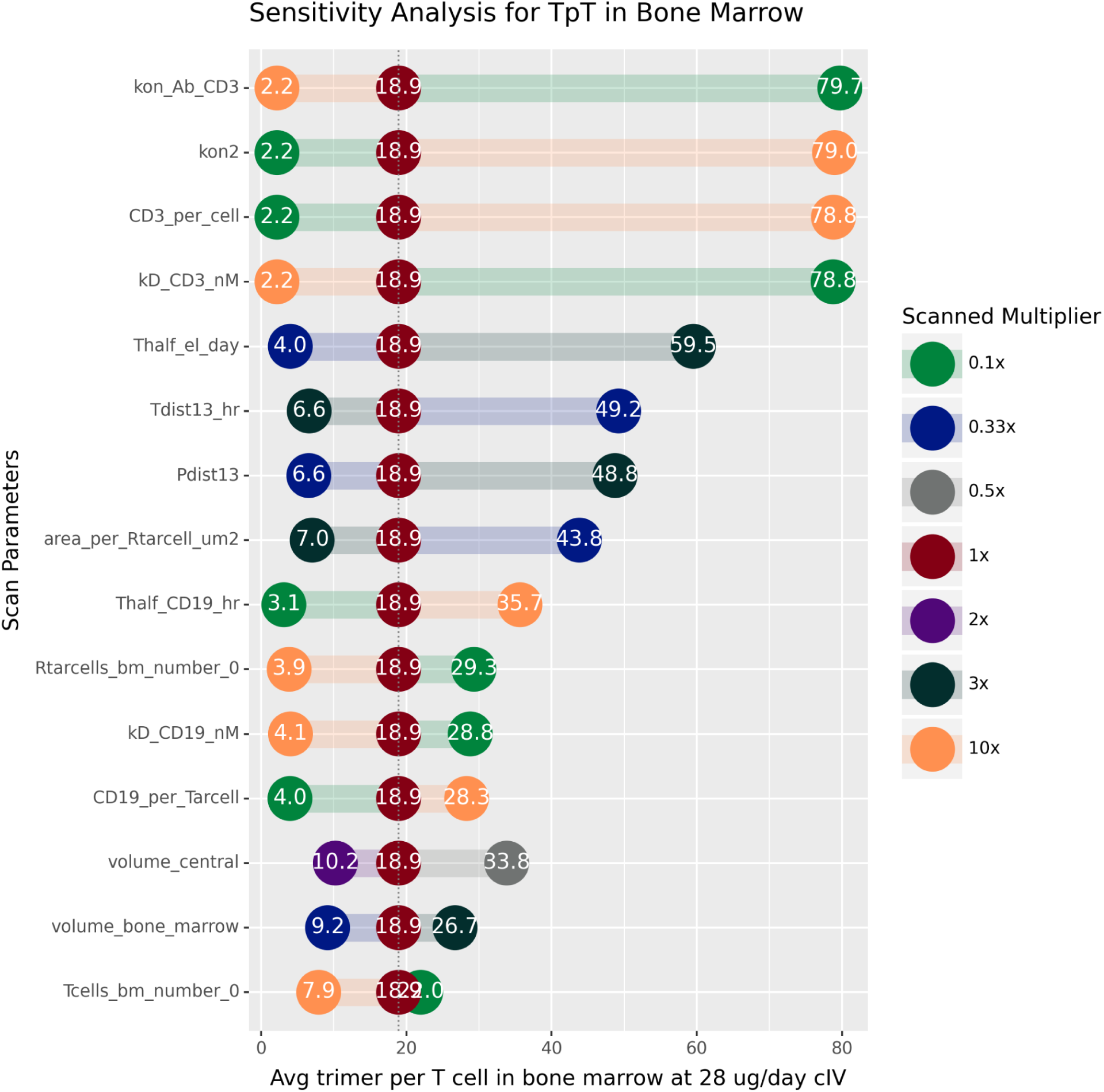
Results from sensitivity analysis with cIV dosing in B-ALL. The dosing schedule was fixed at 28 ug/day. The parameters were varied one at a time by several fold, as indicated in the legend, and the average TpT at day 28 was recorded. The parameters are in rank order based on the highest absolute difference in the results between the top and bottom parameter value. Only the top fifteen values are shown. A description of the parameters and their nominal values are in <u>Table S1</u>.

**Model reaction file. Parameters are taken from <u>Tables S1</u> and <u>S2</u>.**

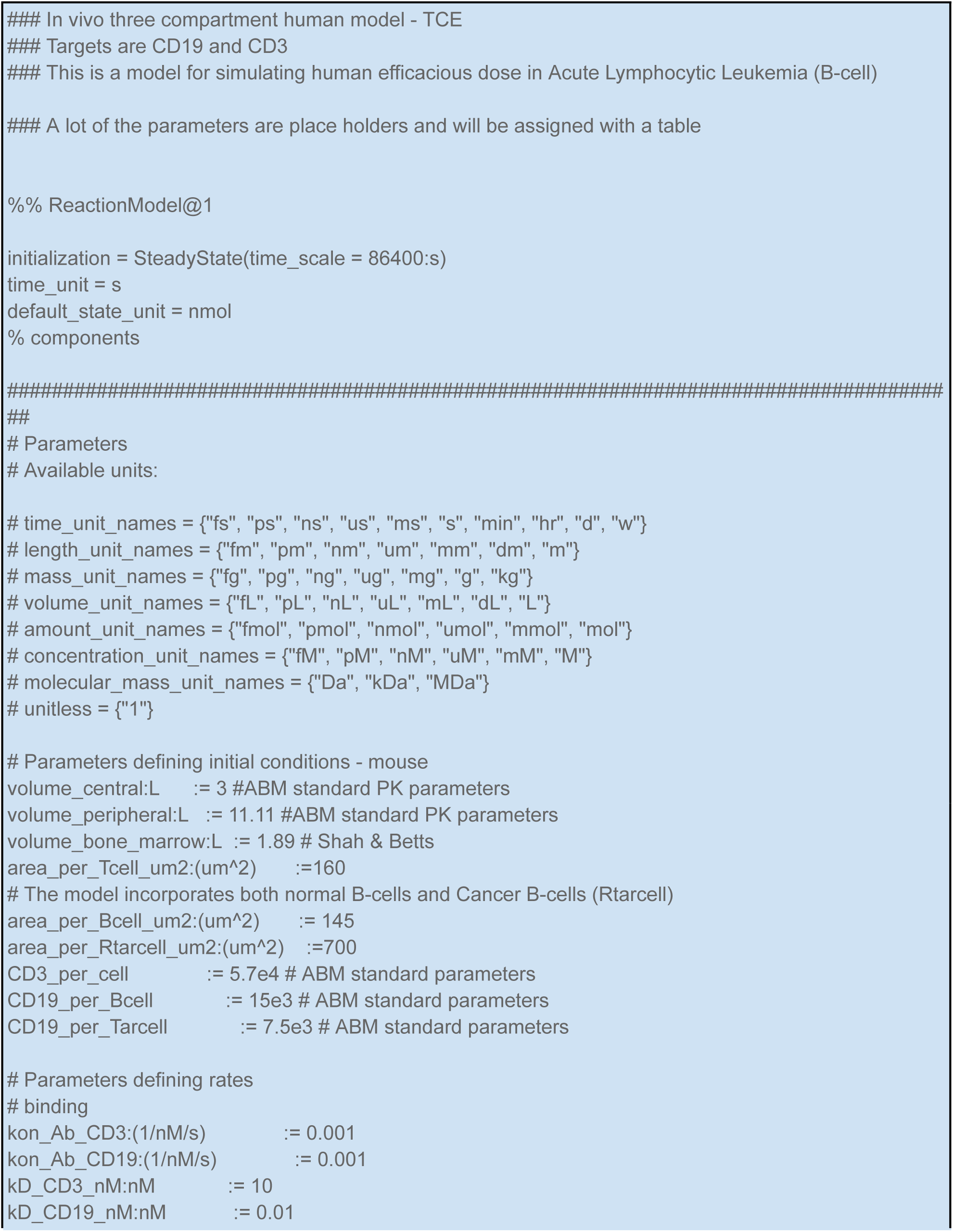
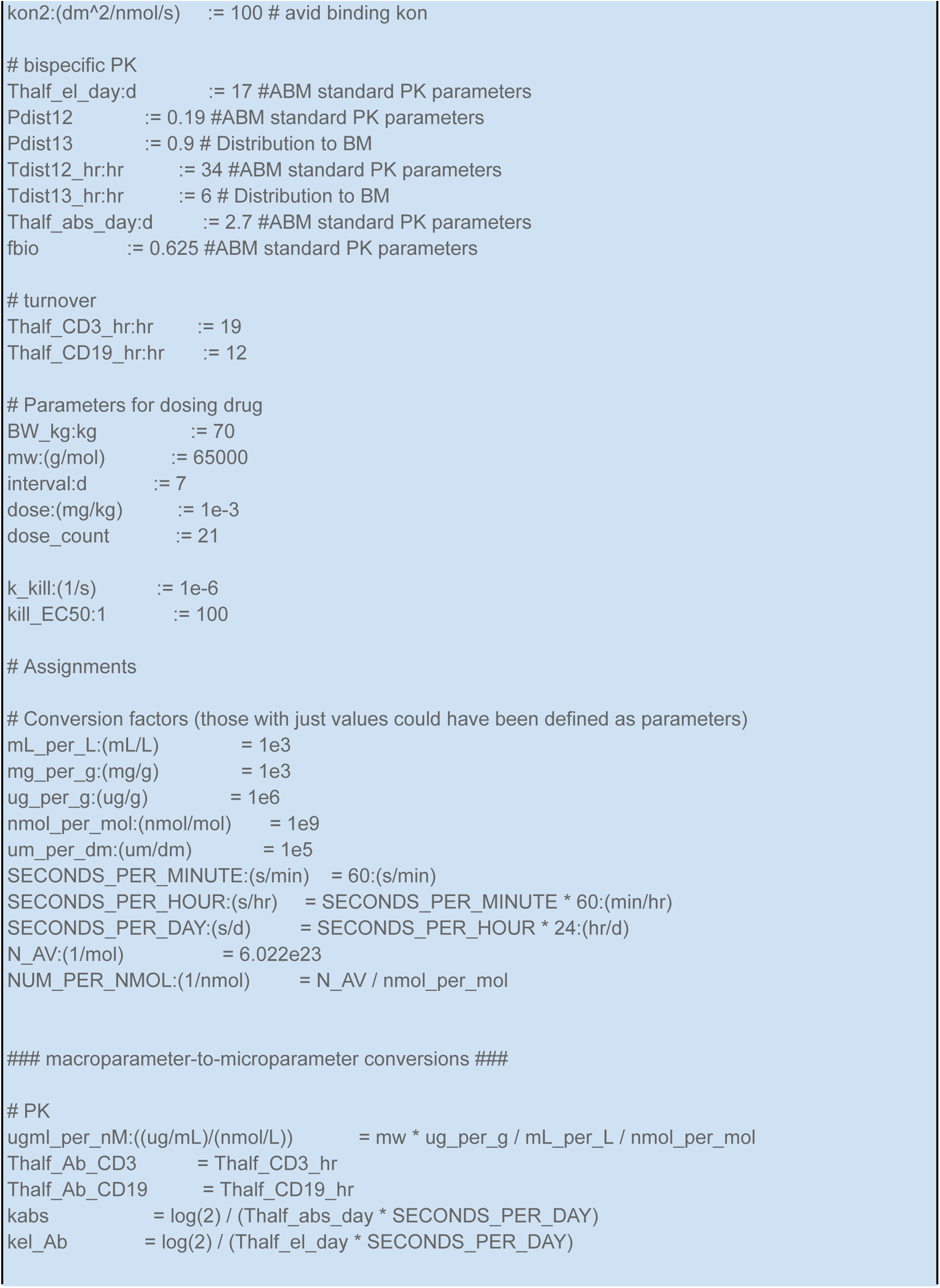
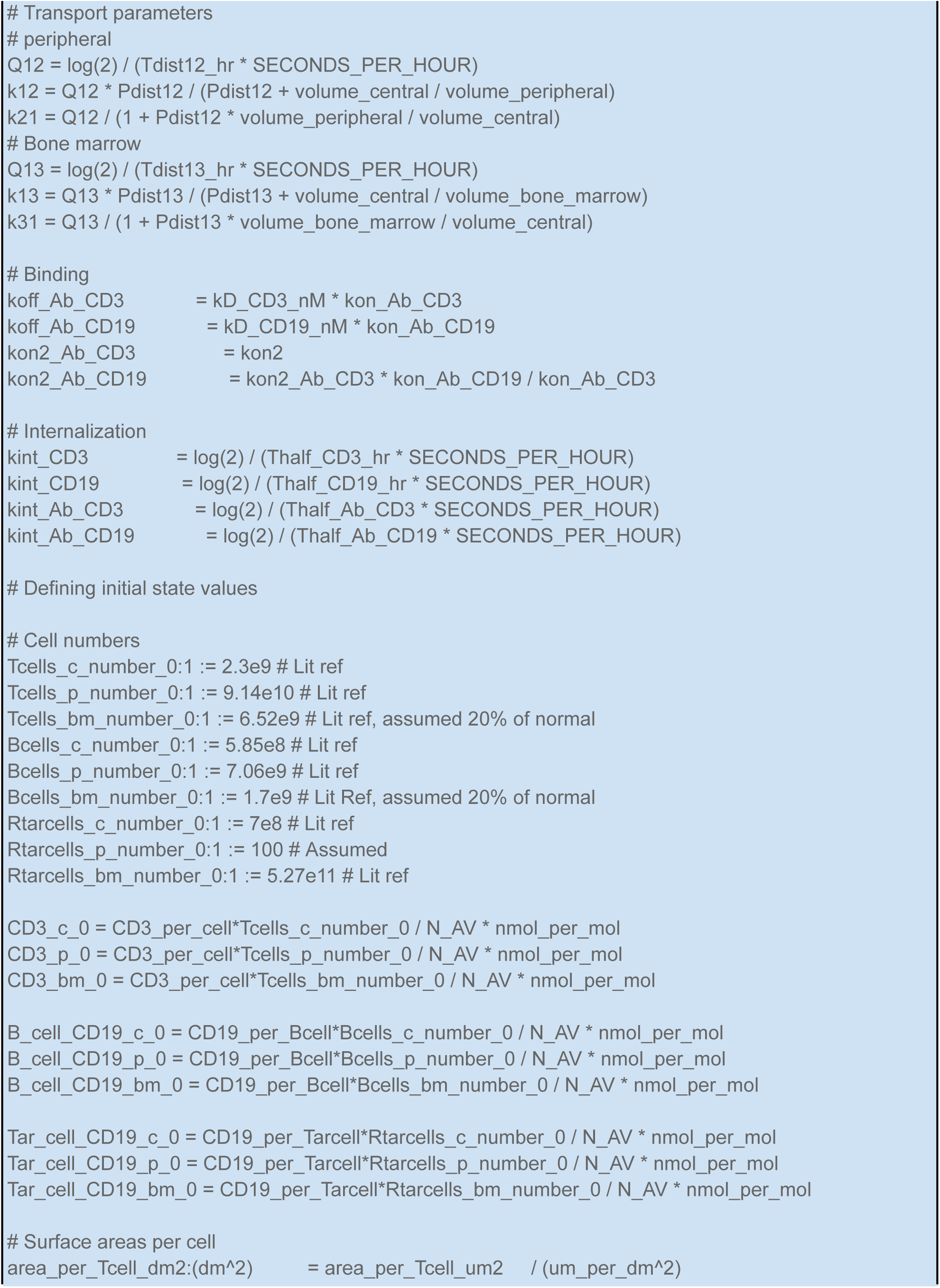
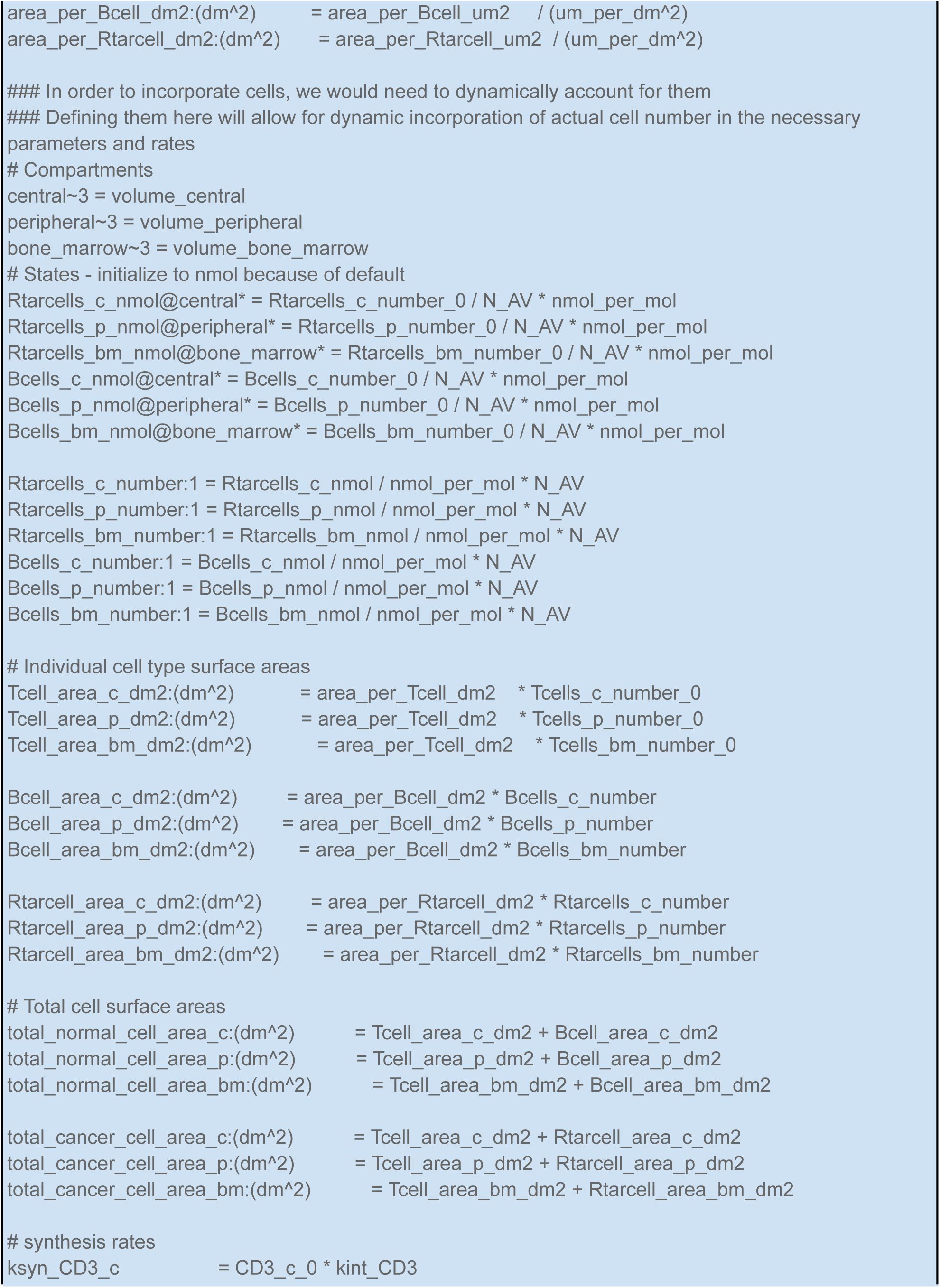
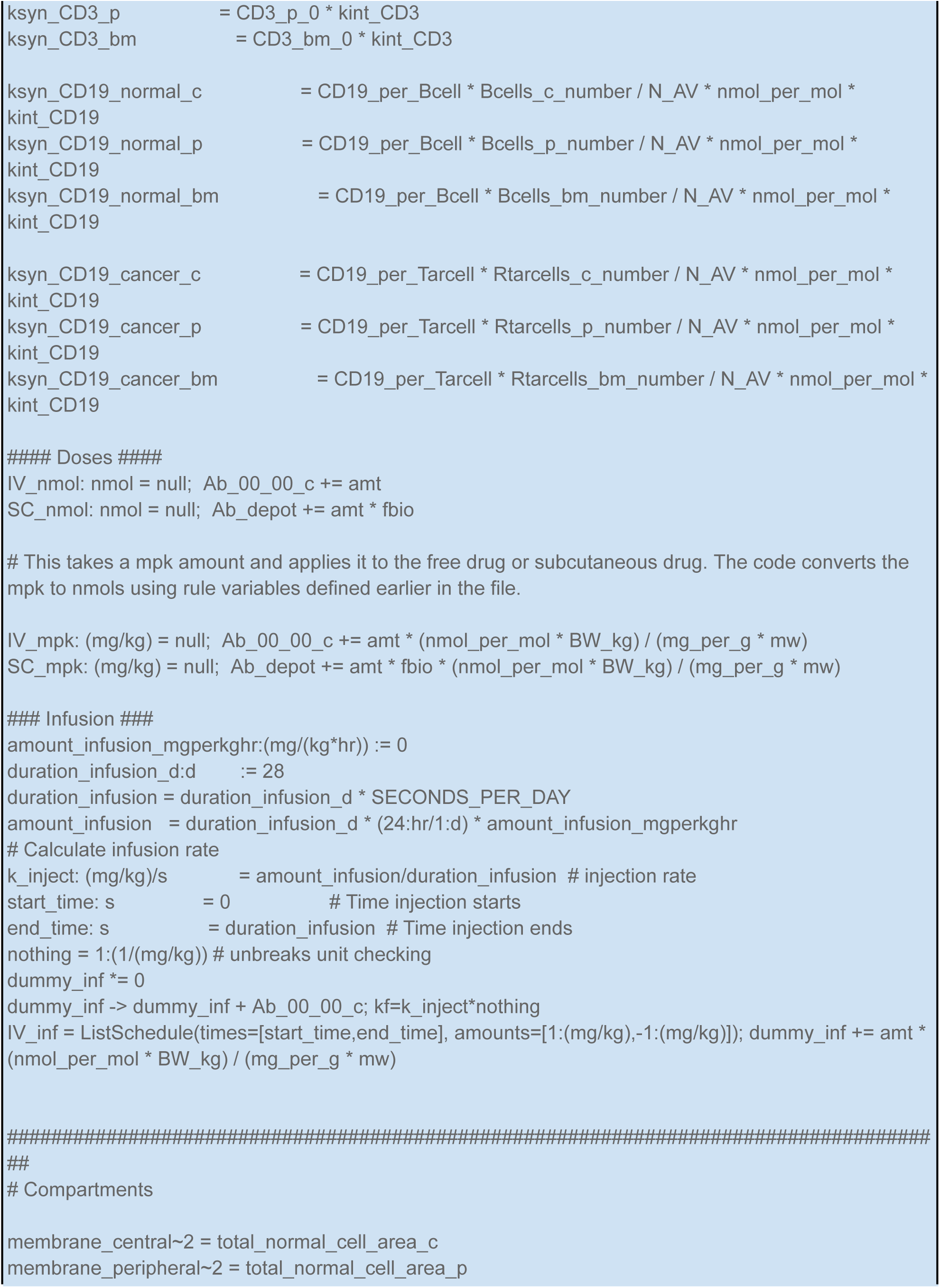
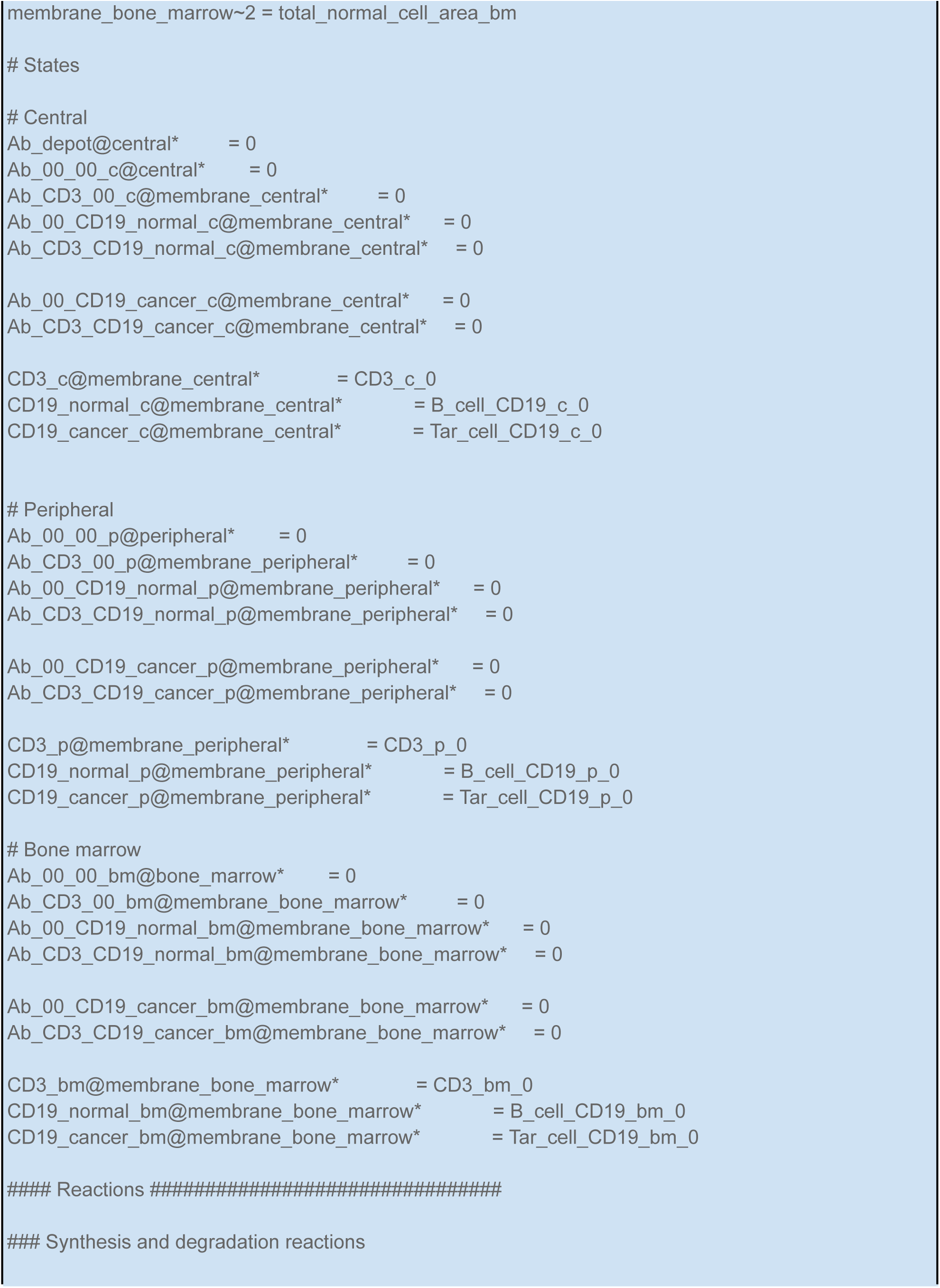
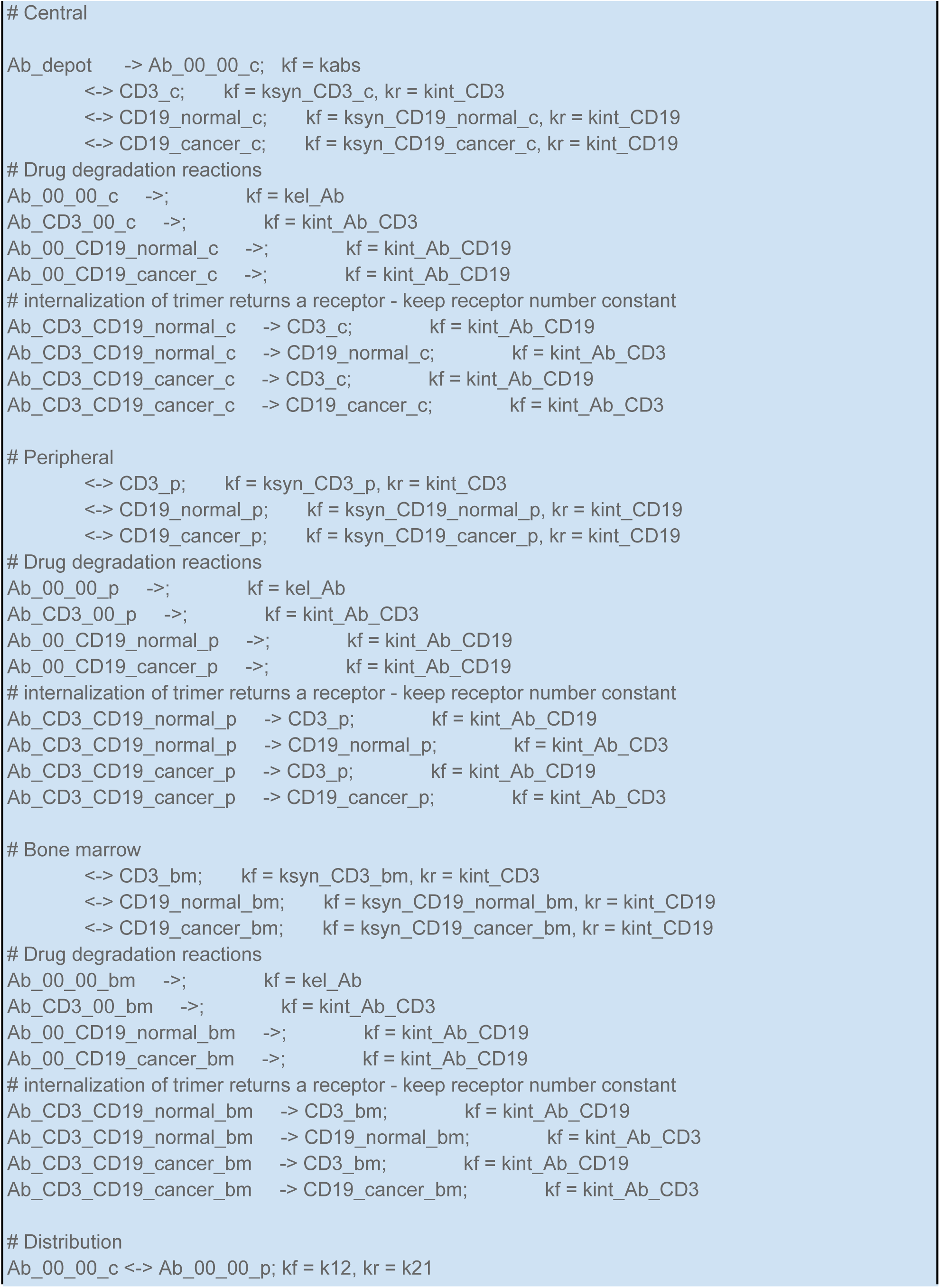
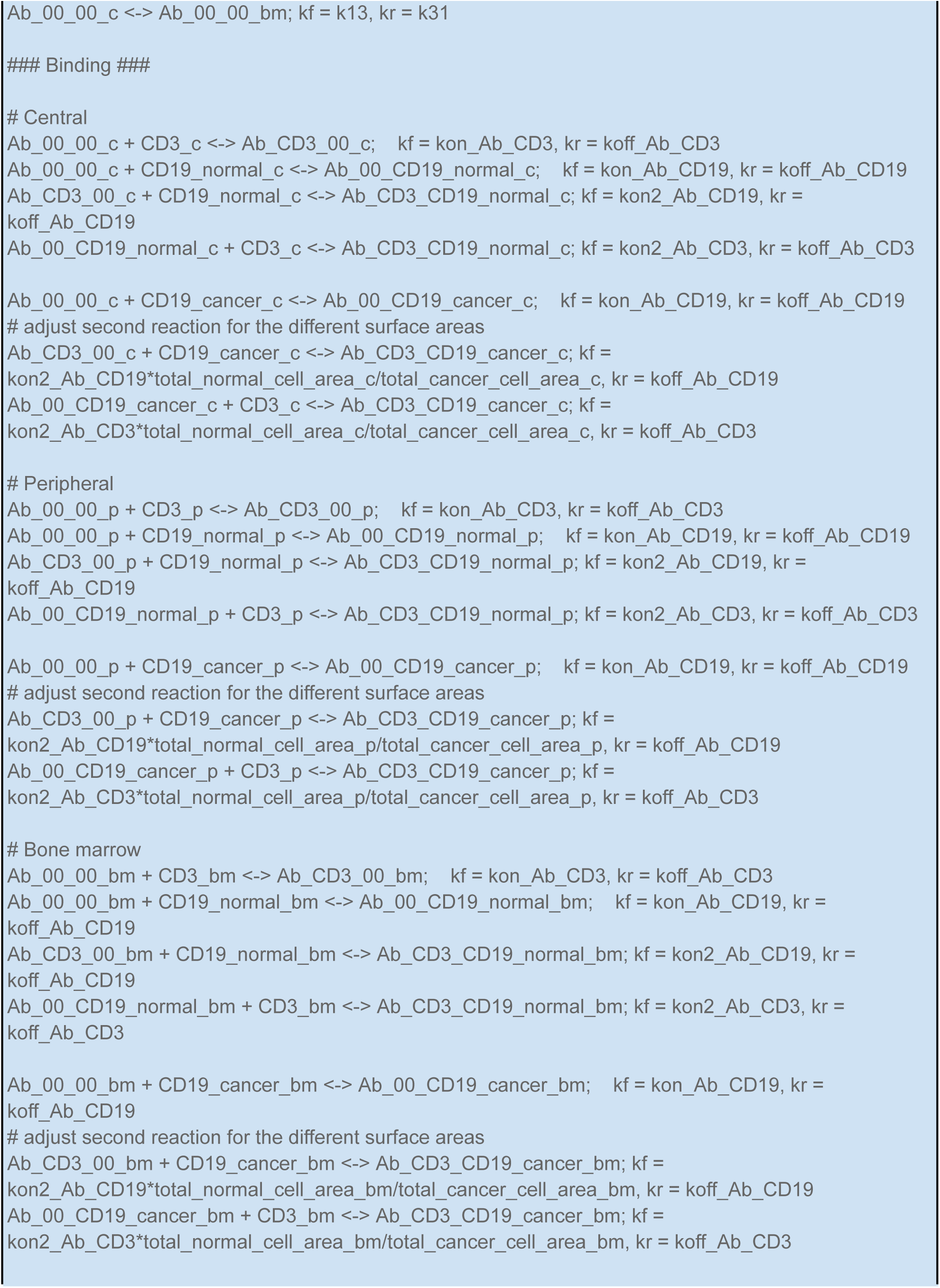
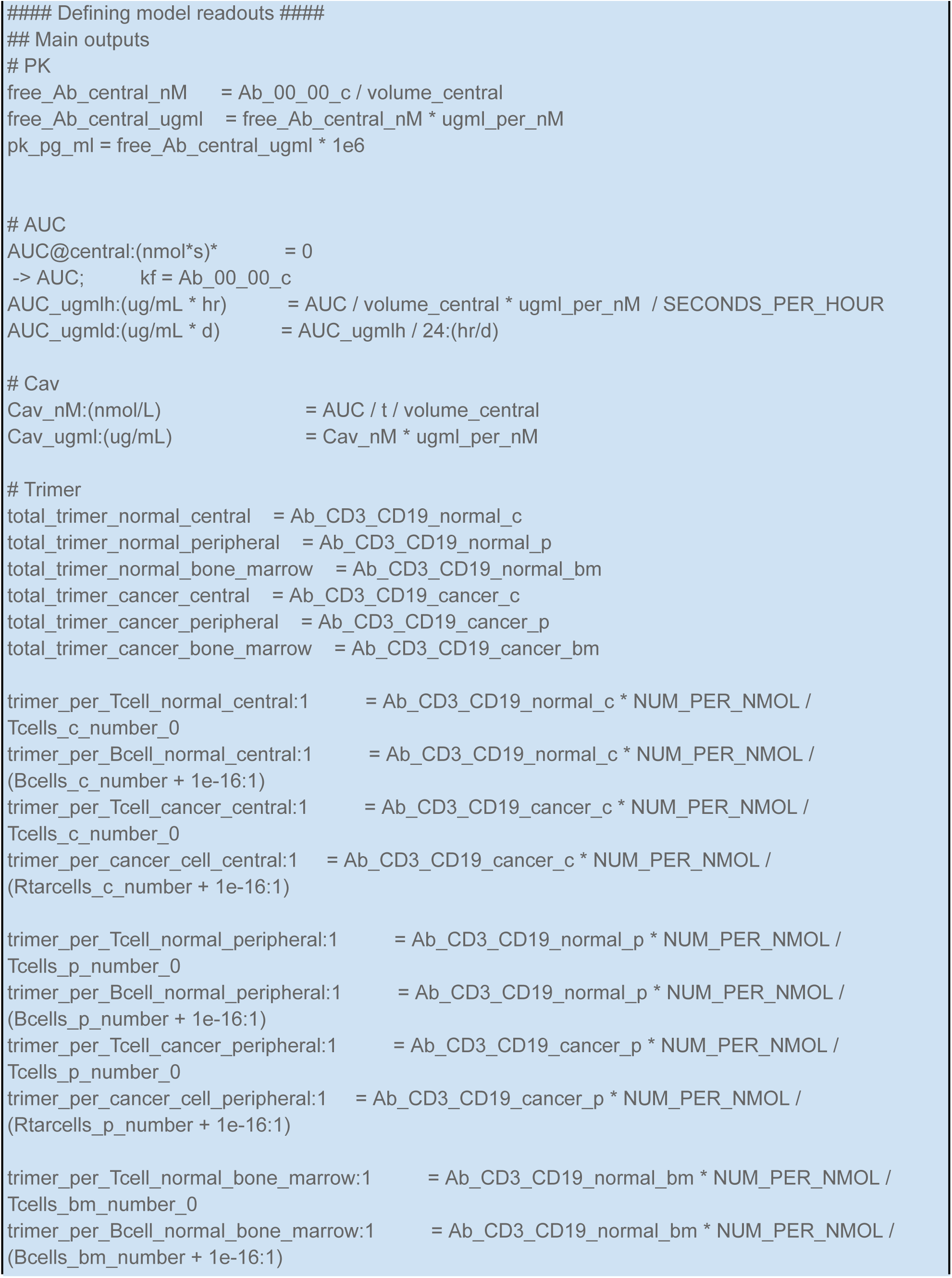
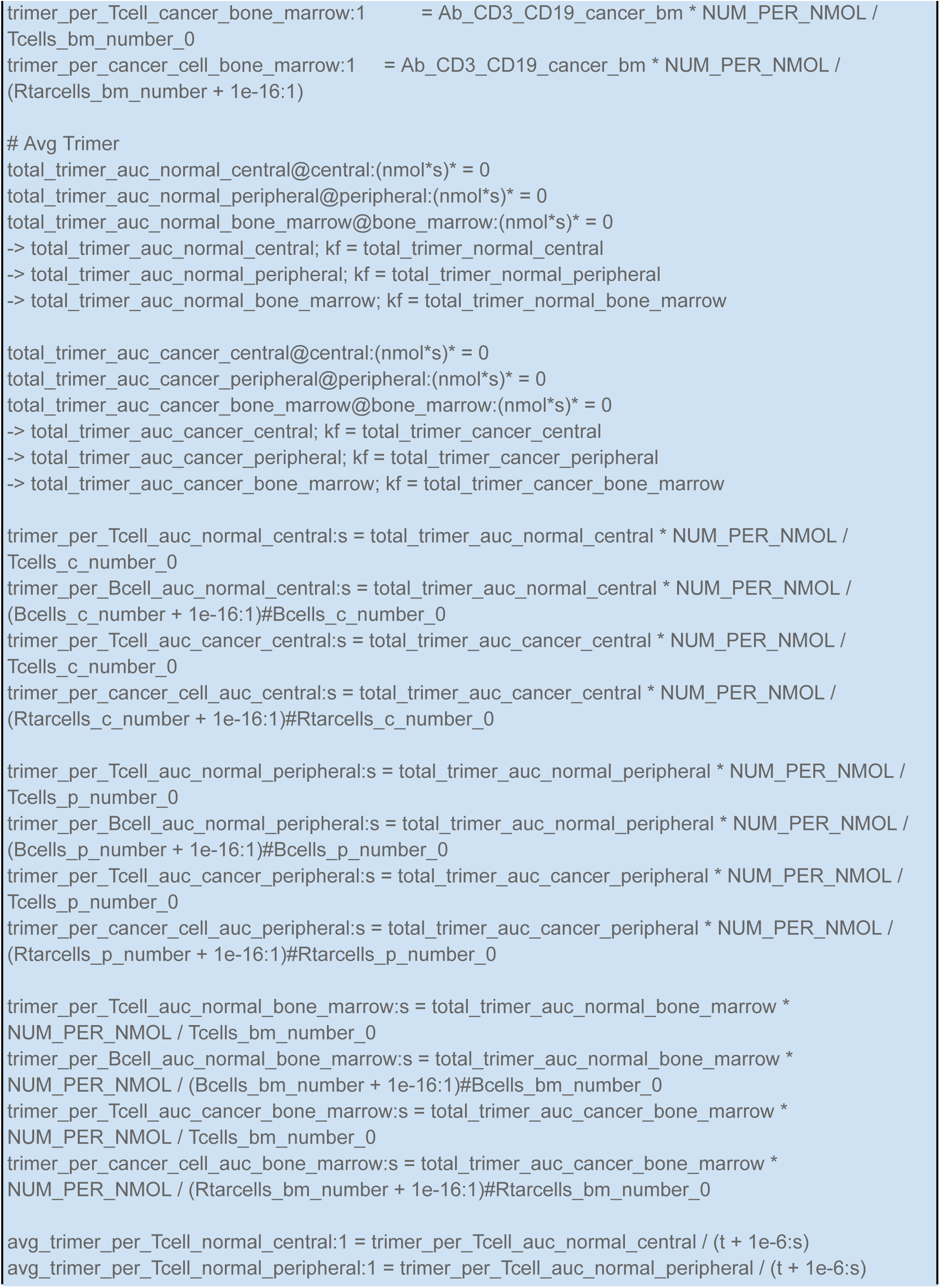
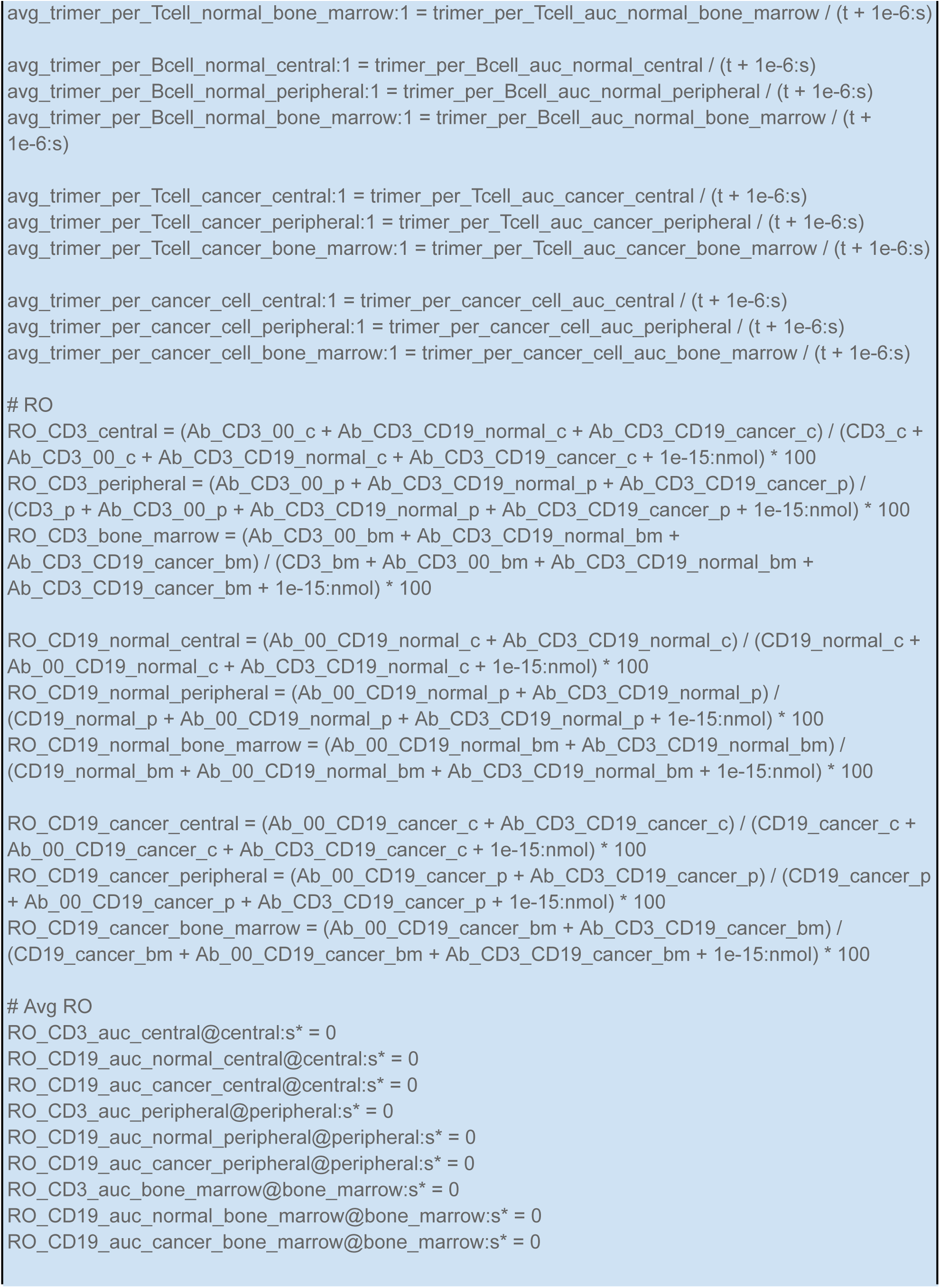
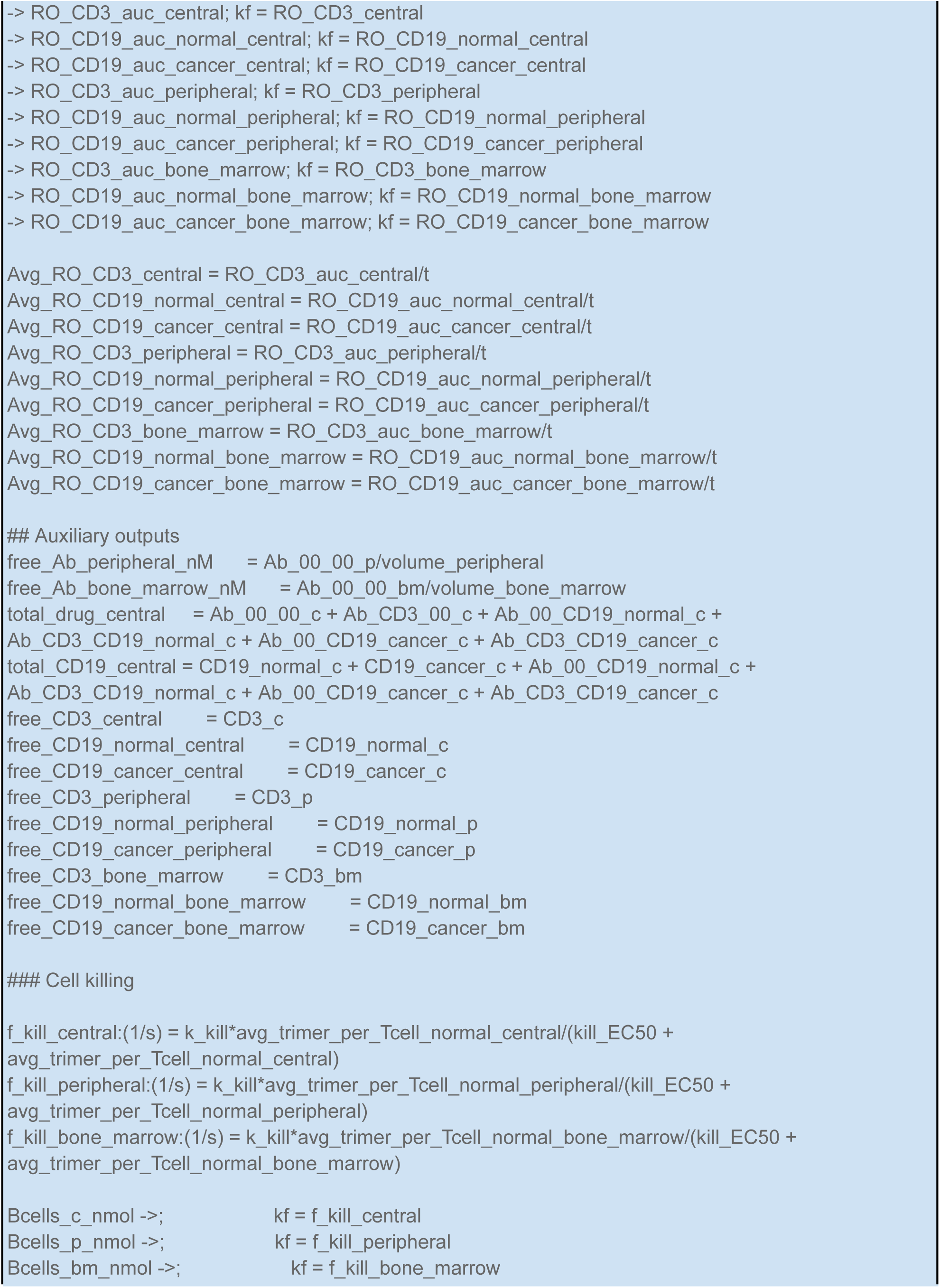
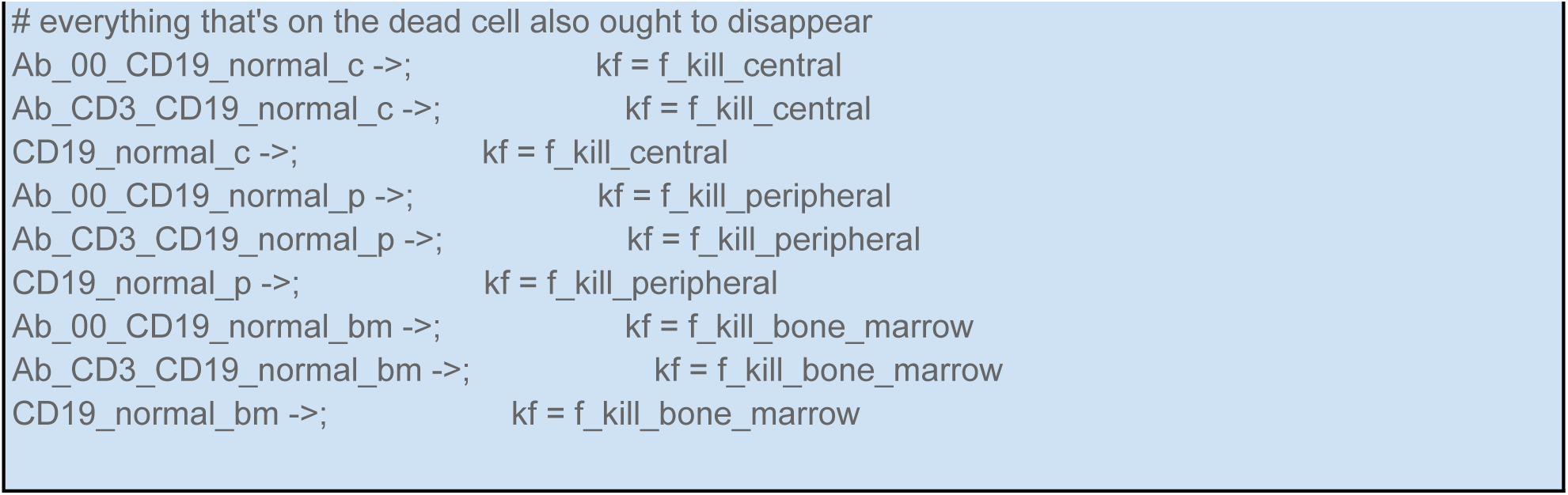

